# Reweighting the UK Biobank to reflect its underlying sampling population substantially reduces pervasive selection bias due to volunteering

**DOI:** 10.1101/2022.05.16.22275048

**Authors:** Sjoerd van Alten, Benjamin W. Domingue, Titus Galama, Andries T. Marees

## Abstract

The UK Biobank (UKB) is a large cohort study of considerable empirical importance to fields such as medicine, epidemiology, statistical genetics, and the social sciences, due to its very large size (∼ 500,000 individuals) and its wide availability of variables. However, the UKB is not representative of its underlying sampling population. Selection bias due to volunteering (*volunteer bias*) is a known source of confounding. Individuals entering the UKB are more likely to be older, to be female, and of higher socioeconomic status. Using representative microdata from the UK Census as a reference, we document significant bias in estimated associations due to non-random selection into the UKB. For some associations, volunteer bias in the UKB is so severe that estimates have the opposite sign. E.g., older individuals in the UKB tend to be in better health. To aid researchers in correcting for volunteer bias in the UKB, we construct inverse probability weights based on UK census microdata. The use of these weights in weighted regressions reduces 78% of volunteer bias on average. Our inverse probability weights will be made available.

## 1 Introduction

In the last decade, several large-scale cohort studies (with N*>*100,000) have been released, collecting a wide range of health-related data from their respondents. These include the UK Biobank (UKB), Lifelines, and the 45 and Up study. Although large in sample size, these studies are not representative of their underlying sampling population [1–5], as they typically relied on respondents to participate voluntarily. Low participation rates (e.g., 5.5% for UKB, 10% for Lifelines, 18% for 45 and Up [6]) result in data sets that are potentially severely selected. These studies are often characterized by “healthy volunteer bias”: typically, study respondents are healthier and of higher socioeconomic status than the population from which they were sampled [4, 7, 8]. For example, on average, UKB respondents are older, more likely to be female, and reside in less socioeconomically deprived areas, compared to the UKB’s sampling population [4, 8].

Such large observational cohort studies are a key resource for medical, epidemiological, statistical genetic, health and social scientific research. The UKB alone has resulted in over 2,500 peer-reviewed publications since its release in 2012 [9]. The lack of representativeness of these biobanks has sparked debate among researchers about the consequences of (possibly unknown) sample selection criteria. Some hold that “[r]egardless of participation rates, as long as there are sufficiently large numbers of participants with different levels of the relevant risk factors under investigation, generalizable associations between baseline characteristics and subsequent health outcomes can be made.”[1] Some have taken this claim so far as to suggest that “representativeness should be avoided” [10, 11]. However, even small deviations from representativeness can cause bias, not just in descriptive statistics (e.g., estimates of disease prevalence), but also in associations between variables of interest [2, 12, 13].

To illustrate the consequences of non-random selection into a non-representative cohort study such as the UKB, consider the simple case where the effect of an exposure *X* on an outcome *Y* is assessed using bivariate linear regression. Figure 1 shows simulations of an exposure *X* ∼ 𝒩 (0, 1) and an outcome *Y* = *X* + *ϵ, ϵ* ∼ 𝒩 (0, 1), and three scenarios (1, 2a and 2b). *X* and *Y* are positively related in the population (the orange and blue dots combined) with slope 1. This is reflected by the orange regression lines in each of the three scatter plots. In scenario 1, individuals with higher values of *Y*, here modeled by a threshold *Y > Y* ^*^, select into the sample (*S*; the blue points) and there is no selection based on *X*. As a result, the regression line estimated within the selected sample *S* (the blue line) is attenuated towards the null. This attenuation bias occurs irrespective of the sign between *X* and *Y*. In scenario 2a, individuals with higher values of *Y* and higher values of *X*, here modeled by a threshold 0.5*Y* + 0.5*X > Z*^*^, select into the sample S. As a result, the regression line estimated within the selected sample *S* is biased downwards. Given the parameters of our simulation, the downward bias is so severe that the regression line is of the incorrect sign. In scenario 2b, individuals with higher values of *Y*, but lower values of *X*, select into the sample *S* (here modeled by a threshold *Y* − 2*X > Z*^*^). Now, the bias is upwards, and the effect of *X* on *Y* is overestimated. Further, when *X* and *Y* both correlate with the probability of selection into *S*, volunteer bias can be introduced even when *X* and *Y* are not related to one another in the underlying sampling population, introducing false positive associations within the sample. Note further that, in all these scenarios, the standard deviations of *X* and *Y* estimated within the selected sample S are smaller, compared to the standard deviations in the full population, as a consequence of selection (this can be seen from the distributions of the blue dots in the simulations, which are more narrow).

**Figure 1:**
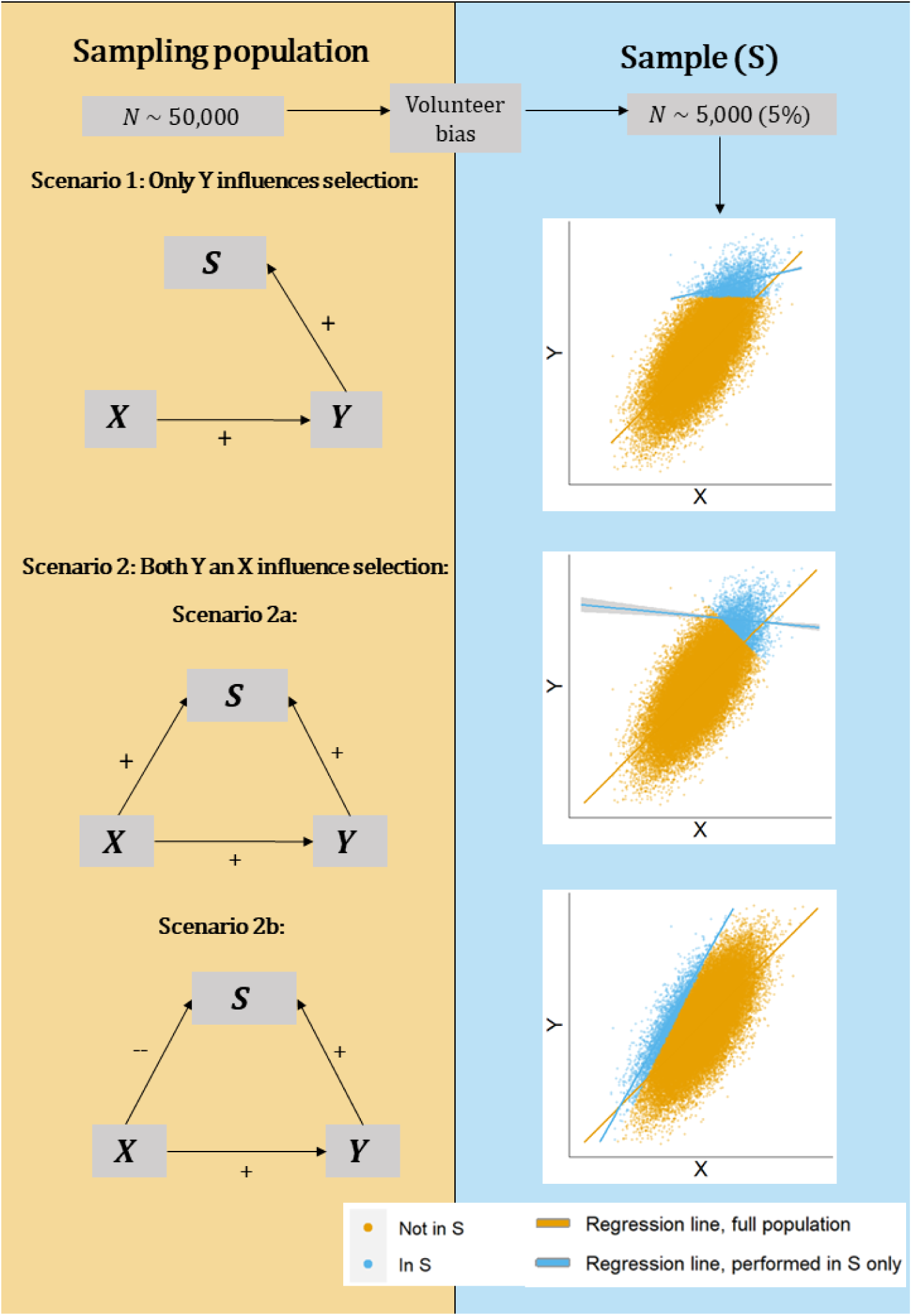
A simulated example of spurious associations due to volunteer bias in a selected sample *S*. In this example, we simulate an exposure *X* ∼ 𝒩 (0, 1) and an outcome *Y* = *X* + *ϵ, ϵ* ∼ 𝒩 (0, 1). *X* and *Y* are positively related in the population (the orange and blue dots combined) with slope 1. This is reflected by the orange regression lines in each of the three scatter plots. In scenario 1, individuals with higher values of *Y*, here modeled by a threshold *Y > Y* ^*\**^, select into the sample (*S*; the blue points) and there is no selection based on *X*. As a result, the regression line estimated within the selected sample *S* (the blue line) is attenuated towards the null. This attenuation bias occurs irrespective of the sign between *X* and *Y*. In scenario 2a, individuals with higher values of *Y* and with higher values of *X*, here modeled by a threshold 0.5*Y* + 0.5*X > Z*^*\**^, select into the sample S. As a result, the regression is downwards biased. Given the parameters of our simulation, the downward bias is so severe that the regression line is of the incorrect sign. In scenario 2b, individuals with higher values of *Y*, but lower values of *X*, select into the sample S (here modeled by a threshold *Y* − 2*X > Z*^*\**^). Now, the bias is upwards, and the effect of *X* on *Y* is overestimated.

It has become apparent that neglecting sample selection into the UKB can lead to misleading estimates. For example, all-cause mortality in the UKB at age 70-74 is about half that of the UK population [4]. Further, effects of risk factors on mortality in the UKB differ markedly from those estimated using more representative data sources [8]. Other work attributes a spurious protective effect of alcohol use on cardiovascular-related mortality to non-random selection into the UKB [14]. Last, volunteer bias is present in genetic associations estimated in the UKB. For example, there are no known biological mechanisms through which genetic markers on the autosome (the “non-sex” chromosomes 1-22) could influence one’s sex. Nonetheless, multiple genetic markers, such as an allele at the FTO gene (on chromosome 16), spuriously associate with the likelihood of being male, a statistical artifact which can be attributed to sex-differential volunteer bias [15].

In this paper, we document the extent to which volunteer bias affects estimated associations in the UKB and attempt to correct for such bias by modeling the selection process. Using a subsample of the UK Census, which we restrict such that it is representative of the population from which the UKB sampled its respondents, and UKB data, we (i) describe how voluntary participation in the UKB results in a data set that is highly non-representative of its underlying sampling population, (ii) show that volunteer bias affects various association statistics of interest in the UKB, and (iii) estimate *inverse probability weights* (IP weights) to estimate association statistics that are representative of the UKB’s sampling population, and thus unaffected by volunteer bias. By comparing weighted associations estimated in the UKB with associations estimated using UK Census data, we infer that the use of our IP weights reduces volunteer bias in the UKB by 78% on average.

## 2 Results

### 2.1 Data and analysis

The goal of the inverse probability weighting (IP weighting) procedure is to make the UKB representative of the *UKB-eligible population* (i.e., all individuals who received an invitation to participate in the UKB). The UKB-eligible population differs from the full population of the United Kingdom in two important aspects. First, the age range is restricted as the UKB only sampled individuals aged between 40 and 69. Second, the geographic range is restricted as only individuals who lived close to any of 22 assessment centers were sampled. These assessment centers were mostly located in urban areas. As Figure 2 shows, this led to a highly uneven sampling of regions, and left out large swaths of Great Britain’s land area and population.^1^

**Figure 2:**
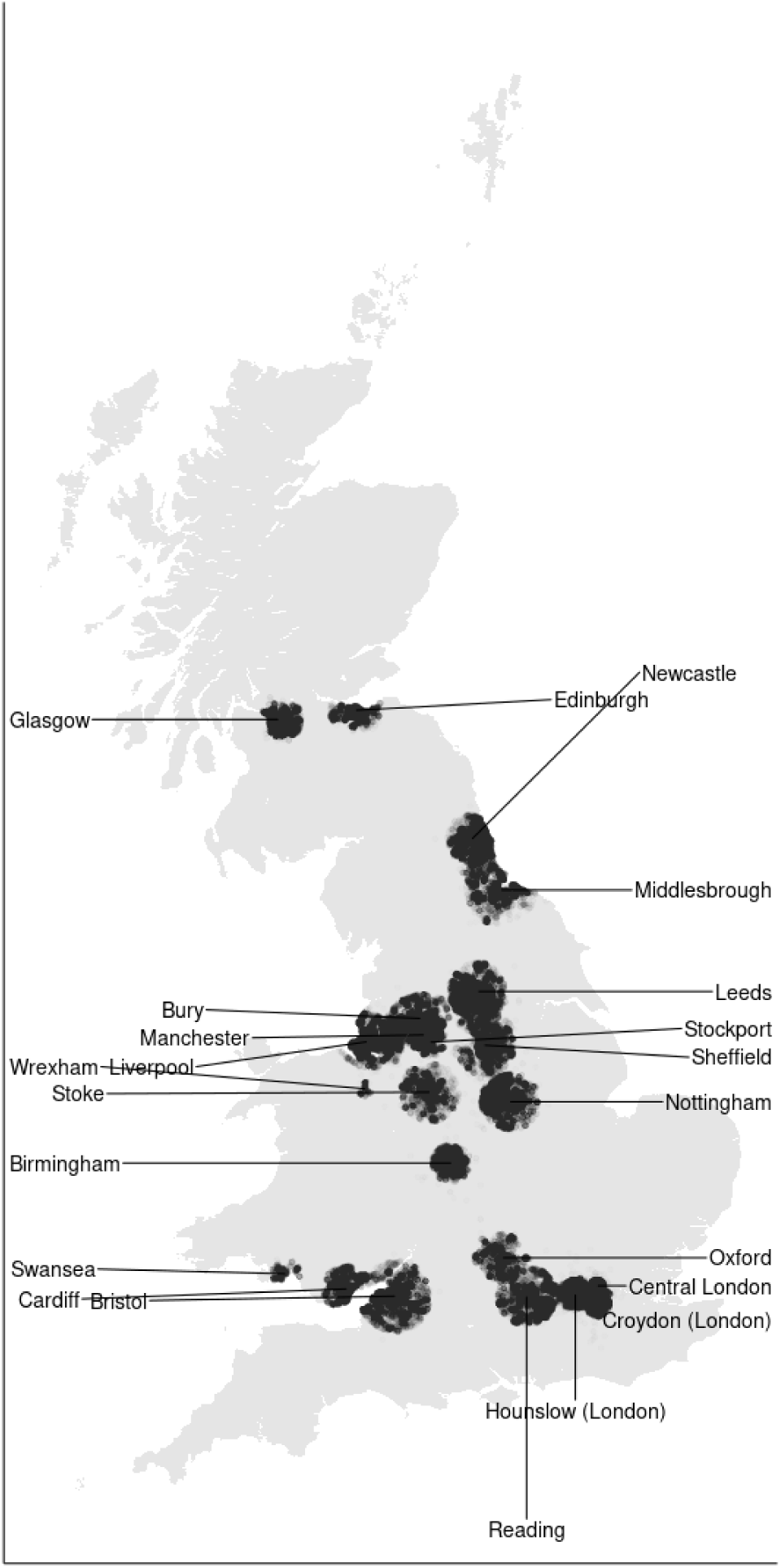
UKB respondent’s location of residence at assessment day. Each black dot corresponds to a UKB respondent in our sample. Only respondents who lived near any of the UKB assessment centers, which were predominantly located in urban areas, received an invitation to participate in the UKB.

To obtain a Census subsample that is representative of this UKB-eligible population, we restrict and weight respondents in the UK Census microdata according to their birth cohort and region of residence, using information on the sampling radii around the 22 assessment centers from where UKB respondents were sampled (See Methods, Section 4). Supplementary Table S1 provides summary statistics for the birth cohorts that were relevant to the UKB (those aged between 40 and 74 at the time of the UK Census), and compares various survey measures for individuals living in Great Britain as a whole *and* the UKB-eligible population. All measures in this table are based on Census data. Compared to the full population, the UKB-eligible population is more ethnically diverse, younger, of lower socioeconomic status (as measured by an overall deprivation indicator), more urbanized (as measured by various proxies for urbanicity), and in worse health. Nonetheless, as we will show in subsequent sections, these differences are relatively small compared to the differences induced by participation in the UKB. The consequences of differences in participation (volunteering) for the UKB are therefore the main focus of this paper.

We use the UKB-eligible subsample of the UK Census microdata to illustrate how the UKB-eligible population differs substantially from UKB respondents in various dimensions. Next, we use this UKB-eligible Census subsample *and* the UKB to estimate IP weights, which are inversely proportional to the likelihood of participation for each UKB respondent (conditional on being within the age range and residing in the area around an assessment center that received invitations).^2^

To estimate IP weights for UKB respondents, we first model the probability of participation in the UKB. We estimate a probit model on UKB data (UKB=1), and our UKB-eligible Census subsample (UKB=0). To predict the UKB participation decision, we use predictors based on year of birth (5-year birth-cohort groups born between 1936 and 1970), sex, ethnicity (White, Mixed, Asian, Black, Other), educational attainment (None, Lower secondary, A-levels/vocational, University), employment status (Paid employment, Retired, Stay-at-home, Incapacitated, Unemployed, Student), region of residence (143 grouped local authorities/grouped council areas that have at least 120,000 inhabitants), tenure of dwelling (Owns house without mortgage, Owns house with mortgage, Shared ownership, Rent, Rent-free), number of cars in the household (0, 1, 2, 3, 4 or more), self-reported health (Bad, Fair, Good/Very Good), and whether the individual lives in a one-person household.

These variables were selected based on several inclusion criteria. First, they had to be assessed for all UKB baseline respondents and UK Census respondents in this age range. Second, they had to be assessed using the same (or very similar) wording in their respective questionnaires. For variables for which the UKB and UK Census categorization was not identical, we harmonized all responses into categories that are comparable in both data sets (see Supplementary Note S1 for detail). When this was not possible, we did not include the variable. Supplementary Table S2 shows the distributions of these variables in the UKB-eligible Census sample and the UKB. For all variables that the data sets have in common, the distributions differ significantly (*p <* 10^−8^) — based on a *χ*^2^-test for equal distributions for each variable — a first indication that volunteer bias affects these variables in the UKB.

All selected variables are either binary or categorical. We enter them non-parametrically in the model by creating a dummy variable for each level the variable can take so that all categorical variables also enter as dummy variables. Further, we include all possible two-way interactions between these dummy variables. As such, our probit model uses 4,820 predictors in its estimation.^3^ To prevent overfitting, we perform variable selection by LASSO estimation (see Methods, Subsection 4.2). Some Census or UKB respondents had one or more variables missing due to item non-response or because some household-level variables in the Census were not assessed for people living in communal establishments. We use an exact matching procedure to impute missing variables (see Supplementary Note S2). After said restrictions and data imputation, the UKB and UK Census data is split into a training sample for estimation of the model for UKB selection (*N*_*UKB*_=98,253, *N*_*UKCensus*_=549,992), and a holdout sample for subsequent testing of the extent to which the estimated weights reduce volunteer bias (*N*_*UKB*_=393,015, *N*_*UKCensus*_=137,499). We deliberately hold out a large part (80%) of the UKB as a testing sample such that weighted analyses can be performed on the vast majority of UKB respondents. By contrast, we include a relatively larger part of the UK Census data (80%) in the training sample, such that the participation probabilities of UKB respondents can be estimated more precisely. To ensure consistency between the training and testing sample, all UKB respondents in the training sample are assigned a weight of 4 in the LASSO probit model estimation. More detail on our methods can be found in Methods (Section 4).

Figure S1 provides a first indication of volunteering bias in the UKB. The UKB participation rate (percentage of those receiving and invitation that actually participate) is 1) low and 2) differs substantially by region (ranging between 2.83% at the 25th and 5.81% at the 75th percentile of the distribution).

### 2.2 Evidence for volunteer bias in UKB

A comparison of the UKB and the UKB-eligible subsample of the UK Census reveals substantial non-random selection of UKB participants from the UKB-eligible population (Table 1). Compared to the UKB-eligible (invited) population, individuals who participated in the UKB are older, healthier, higher educated, of higher socioeconomic status, and more likely to be white. For all variables included in the table, we find that their means differ significantly between the UKB and the UKB-eligible population (*p <* 10^−8^ for all variables included in the table). Some differences are quite large. For example, individuals in the UKB-eligible population are over twice as likely to report being in poor health, compared to those who decided to participate in the UKB (9.3 versus 4.4 percent), despite the fact that UKB participants are ∼ 3.5 years older on average. Individuals with low socioeconomic status are similarly underrepresented in the UKB. This is, for example, apparent in rates of home ownership: 89.9% amongst UKB respondents versus 73.6% in the UKB-eligible population.

**Table 1:** Summary statistics for the UKB-eligible population (based on 2011 UK Census data), the UKB, and the weighted UKB. Summary statistics are based on the combined training and holdout samples for the UKB and UK Census data. For all variables shown here, mean values differ significantly between the UKB-eligible population (rows indicated by “Census (UKB-eligible)”) and the UKB (*P <* 10^*-*8^). Observations from the rows indicated by “UKB Weighted” show summary statistics for the UKB sample after applying IP weighting.

| Var | Dataset | N | Min | Mean | Max | SD |
| --- | --- | --- | --- | --- | --- | --- |
| **Discrete/Continuous variables** |  |  |  |  |  |  |
| Year of Birth | Census (UKB-eligible) | 687,491 | 1938 | 1955.101 | 1968 | 9.039 |
|  | UKB | 491,268 | 1938 | 1951.545 | 1968 | 8.261 |
|  | UKB Weighted | 491,240 | 1938 | 1954.728 | 1968 | 8.905 |
| Self-reported health | Census (UKB-eligible) | 687,489 | 1 | 2.625 | 3 | 0.648 |
|  | UKB | 488,956 | 1 | 2.701 | 3 | 0.546 |
|  | UKB Weighted | 488,941 | 1 | 2.644 | 3 | 0.625 |
| Years of education | Census (UKB-eligible) | 687,489 | 7 | 12.608 | 20 | 5.071 |
|  | UKB | 480,251 | 7 | 13.585 | 20 | 4.987 |
|  | UKB Weighted | 480,233 | 7 | 12.925 | 20 | 5.036 |
| No. of cars | Census (UKB-eligible) | 683,138 | 0 | 1.364 | 4 | 0.966 |
|  | UKB | 487,832 | 0 | 1.547 | 4 | 0.870 |
|  | UKB Weighted | 487,820 | 0 | 1.396 | 4 | 0.959 |
| **Bivariate variables** |  |  |  |  |  |  |
| Female | Census (UKB-eligible) | 687,491 | 0 | 0.508 | 1 | 0.500 |
|  | UKB | 491,268 | 0 | 0.546 | 1 | 0.498 |
|  | UKB Weighted | 491,240 | 0 | 0.509 | 1 | 0.500 |
| University or equiv. | Census (UKB-eligible) | 687,489 | 0 | 0.278 | 1 | 0.448 |
|  | UKB | 480,251 | 0 | 0.336 | 1 | 0.472 |
|  | UKB Weighted | 480,233 | 0 | 0.292 | 1 | 0.455 |
| Reports “poor health” | Census (UKB-eligible) | 687,489 | 0 | 0.093 | 1 | 0.290 |
|  | UKB | 488,956 | 0 | 0.044 | 1 | 0.206 |
|  | UKB Weighted | 488,941 | 0 | 0.081 | 1 | 0.273 |
| Has paid work | Census (UKB-eligible) | 687,491 | 0 | 0.609 | 1 | 0.488 |
|  | UKB | 486,711 | 0 | 0.579 | 1 | 0.494 |
|  | UKB Weighted | 486,693 | 0 | 0.615 | 1 | 0.487 |
| Retired | Census (UKB-eligible) | 687,491 | 0 | 0.249 | 1 | 0.432 |
|  | UKB | 486,711 | 0 | 0.341 | 1 | 0.474 |
|  | UKB Weighted | 486,693 | 0 | 0.252 | 1 | 0.434 |
| Incapacitated | Census (UKB-eligible) | 687,491 | 0 | 0.069 | 1 | 0.254 |
|  | UKB | 486,711 | 0 | 0.033 | 1 | 0.179 |
|  | UKB Weighted | 486,693 | 0 | 0.064 | 1 | 0.244 |
| Unemployed | Census (UKB-eligible) | 687,491 | 0 | 0.033 | 1 | 0.179 |
|  | UKB | 486,711 | 0 | 0.016 | 1 | 0.127 |
|  | UKB Weighted | 486,693 | 0 | 0.033 | 1 | 0.178 |
| House owner | Census (UKB-eligible) | 683,138 | 0 | 0.736 | 1 | 0.441 |
|  | UKB | 484,157 | 0 | 0.899 | 1 | 0.302 |
|  | UKB Weighted | 484,147 | 0 | 0.756 | 1 | 0.430 |
| White ethnicity | Census (UKB-eligible) | 687,491 | 0 | 0.888 | 1 | 0.315 |
|  | UKB | 491,268 | 0 | 0.946 | 1 | 0.226 |
|  | UKB Weighted | 491,240 | 0 | 0.893 | 1 | 0.310 |
| One-person household | Census (UKB-eligible) | 683,138 | 0 | 0.181 | 1 | 0.385 |
|  | UKB | 487,922 | 0 | 0.185 | 1 | 0.388 |
|  | UKB Weighted | 487,908 | 0 | 0.188 | 1 | 0.391 |

For all discrete and continuous variables, we also observe smaller standard deviations in the UKB, compared to the UKB-eligible population. Such a decrease in standard deviations across all variables is consistent with non-random sample selection: it implies that these variables have a narrower distribution in the UKB. This is consistent with the notion that individuals of a certain type are more likely to get sampled in the UKB. This pattern of smaller standard deviations is also in line with our simulations as can be visually seen in Figure 1.^4^

Our LASSO probit model adequately discriminates between UKB and UK Census observations, with an area under the curve (AUC) of 0.767 [16] (*IMV* = 0.006 [17]) in the independent holdout sample. For comparison, our AUC is similar to the AUC achieved when predicting mortality in the Health and Retirement Study to correct for mortality selection bias [18]. Figure S2 shows a variable importance plot in which we assess the extent to which the model’s performance in the holdout sample degrades when permuting each variable in the sample, one at a time. Through each permutation, the respective variable becomes unrelated to the model’s outcome, and hence the relative contribution of that variable to the model’s performance can be assessed: the larger the reduction in AUC after leaving the variable out, the more important that variable is to the model’s performance. The variables region, year of birth, and education drive most of the performance of the LASSO model.

We use predictions from the LASSO model to create IP weights for the UKB 80% holdout sample. Figure S3 visualizes the distribution of these weights. To reduce the influence of outliers resulting from UKB participation probabilities that are estimated to be very close to zero, we winsorized the weights by setting values lower than the 1st percentile equal to that of the 1st percentile, and values higher than the 99th percentile equal to the value of the 99th percentile, as is recommended in the literature, [19].

Weighted means of summary statistics estimated in the UKB show that, after IP weighting, the UKB becomes much closer to being representative of the UKB-eligible population (Table 1; compare the rows “UKB” to “UKB-eligible Census” and “UKB Weighted” to “UKB-eligible Census”). As we found for the means, applying IP weights to the UKB also makes the standard deviations much more reflective of the standard deviations in the UKB-eligible population.

Further, we assess how much means of variables *measured in the UKB but not in the UK Census* change after applying our IP weights. Table S3 shows summary statistics of various variables in the UKB sample before and after weighting. Overall, the UKB after weighting is younger, heavier, in worse (mental) health, more likely to smoke, and of lower socioeconomic status, compared to the unweighted UKB sample, which is consistent with healthy volunteer bias. For example, weighting the UKB increases the Townsend deprivation index from -1.317 to -0.43 (0 being the national UK average). This is consistent with the fact that the UKB oversampled respondents from high socioeconomic status areas. We further find that weighting especially increases the prevalence of addictive behaviors in the UKB. For example, the percentage of respondents indicating they were “ever addicted” to any substance or behavior increases from 6% in the UKB to 7.4% in the UKB after IP weighting, a difference of 22.7%. Similarly, the percentage of respondents that ever smoked cannabis increases from 44.2% to 54.3% after IP weighting. Other substantial differences occur for respondents reporting chest pain or discomfort in the chest (16.2% before, and 18.6% after applying IP weighting) and for respondents being breastfed as a child (72.3% before, and 68.4% after weighting). For other variables, for example anthropometric ones, the application of IP weights results in negligible differences.

### 2.3 Volunteer bias in associations estimated in the UKB

We first assess whether volunteer bias affects associations estimated in the UKB. We estimate bivariate linear probability models in the UKB and the UKB-eligible Census population separately for several binary variables that the UKB and UK Census have in common. Figure 3 plots the resulting coefficients (with their width indicating the 95% confidence interval in the estimate) in the UKB-eligible UK Census data (yellow bars) and the UKB (blue bars). P-values for the null hypothesis that the point estimates in the UKB-eligible population and the UKB are the same (see Methods for additional detail, Section 4.3) are shown to the right of each association test. A comparison of blue and yellow bars suggests that volunteer bias severely biases associations between basic demographic variables: for *all* estimated linear probability models, the UKB-eliglible UK Census and UKB point estimates are significantly different from one another. Importantly, the size of these differences is often very large. For example, the association between being employed and reporting poor health amongst UKB respondents (CI_95_ = [-0.306; -0.292]) is substantially weaker than the association in the broader UKB-eligible population, (CI_95_ = [-0.513; -0.498]). The difference between these point estimates is highly statistically significant (*P <* 10^−8^). Estimating these types of models in the UKB can thus result in sizeable distortions of the actual associations in the underlying sampling population.

**Figure 3:**
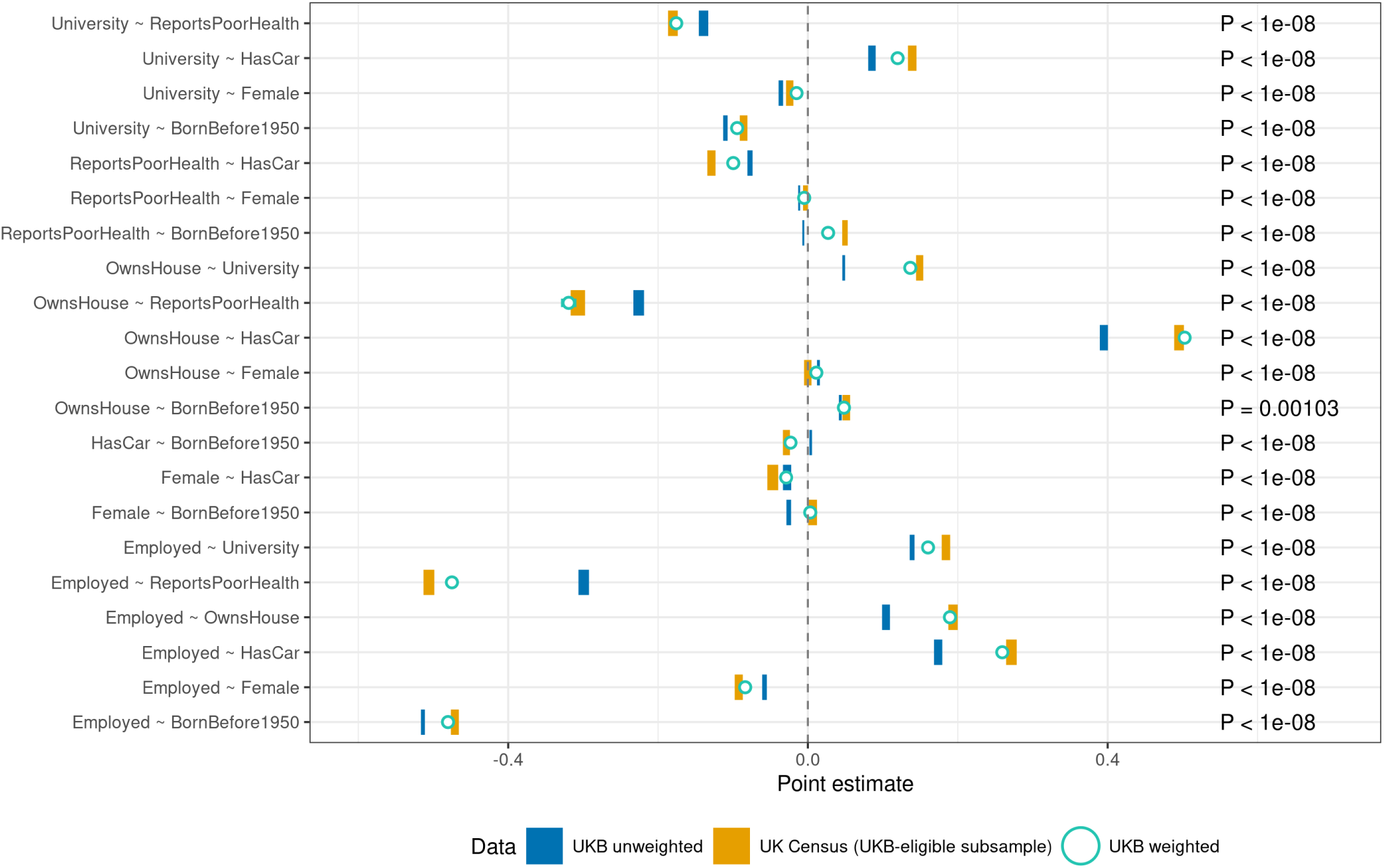
Estimated associations based on bivariate linear probability models in the UKB and the UKB-eligible UK Census data: Each blue bar is estimated in the UKB (80% holdout sample) using OLS. Associations in the UKB-eligible UK Census (yellow bars, 20% holdout sample) are estimated by a weighted least squares (WLS) model using the UK Census adjustment factors to ensure representativeness of the UKB-eligible population (constructed as described in Methods, Section 4.1.3). The green open circles show WLS models estimated in UKB, with weights inversely proportional to selection into the UKB, as estimated from the LASSO probit model that predicts UKB volunteering. 95% confidence intervals are indicated by the width of the bar (heteroskedasticity-robust standard errors). All blue and yellow bars, except for one, are highly significantly different (*P <* 10^*-*8^). IP weighting leads to substantially improved associations, since the green dots are substantially closer to the yellow bars, compared to the blue bars.

We find numerous instances of associations estimated in the UKB that are false positives, or even of the incorrect sign. First, the UKB indicates that individuals born before 1950 are *less* likely (*CI*_95_ = [-0.007; -0.005]) to report being in poor health than younger individuals. While this association is small, it is contrary to the vast evidence that health deteriorates as we age. In the UKB-eligible population, we do observe the expected positive association between reporting being in poor health and being born before 1950 (*CI*_95_ = [0.0458; 0.0531]). Another example of an incorrect sign is that the UKB indicates that women are less likely to be born before 1950 than men (*CI*_95_ = [-0.0288; -0.0226]), whereas the reverse holds in the UKB-eligible population (*CI*_95_ = [0.0004; 0.0122]), which is consistent with the fact that men live shorter than women. Last, young people in the UKB (born before 1950) are slightly more likely to own a car compared to older respondents (*CI*_95_ = [0.002; 0.0055]), whereas the association in the UKB-eligible population is the reverse (*CI*_95_ = [-0.029; -0.033]). We also find an example of a false positive result in the UKB: according to the UKB, women are more likely to own a house than men (*CI*_95_ = [0.012; 0.016]), whereas in the UKB-eligible population, the corresponding point estimate is indistinguishable from zero. All other associations are of the same sign in both the UKB and the UKB-eligible population, but are nonetheless substantially different from one another, such that the UKB may severely overstate or understate the relationships between various variables.

In Figure S5, we show a similar plot, but for bivariate linear models between several discrete and/or continuous variables. Here, we again find that the UKB fails to show that younger people are healthier (for the coefficient on year of birth (*CI*_95_ = [-0.0018; 0.0044] in the UKB, [0.199; 0.21] in the UKB-eligible population). Further, the UKB association incorrectly indicates that women are younger on average than men (*CI*_95_ = [0.021; 0.027] in the UKB, [-0.008; -0.013] in the UKB-eligible population). Last, women in the UKB are in better health (*CI*_95_ = [0.05; 0.057]), whereas the underlying sampling population shows no difference between men and women (*CI*_95_ = [-0.007; 0.004]).

### 2.4 Inverse probability weighting mitigates volunteer bias in associations estimated in the UKB

We next report results for the same models estimated in the UKB, using IP weighted regressions to correct for volunteer bias (see green dots in Figure 3, including 95% confidence intervals). Overall, the associations in the UKB become less biased, i.e., after correction for volunteer bias via IP weighting, the green open circles are substantially closer to the estimates in the “true” UKB-eligible population (yellow bars) than the original unweighted associations (blue bars). Quantifying the size of the bias as the absolute difference between the point estimates in UKB-eligible UK Census data and the UKB, we conclude that the average bias reduction over all models shown in the figure is 78%. In Supplementary Note S3, we show that similar results hold for associations between discrete and/or continuous variables (Figure S5), with an average bias reduction of 77% over all our estimated associations. There, we also discuss that the ability to capture selection bias of the IP weights is robust to different specifications of our LASSO model, such as omitted variables that predict selection, or insufficient harmonization between UKB and UK Census variables (Figure S6, Figure S7).

## 3 Discussion

We have provided several examples of how associations estimated in the UKB could be misleading when volunteer bias is not taken into account. These examples include both false positives and even associations that have the incorrect sign when naively estimated in the UKB. In perhaps the most striking example, the UKB suggests a positive effect of age on self-reported health, whereas the expected negative effect persists in the underlying sampling population. We have further shown that the computation of IP weights for the UKB is feasible, and that the use of such weights greatly reduces volunteer bias in association statistics.

Our analyses highlight the drawbacks of non-random sampling for valid statistical inferences of associations that aim to inform medical, epidemiological, or social-scientific theories, but at the same time also highlights that corrections, using IP weights, can be made. To facilitate the use of our IP weights, we will release these weights as a derived data field to the UKB for use by UKB-approved researchers.

Researchers who work with data sets other than the UKB, that may suffer from similar biases due to volunteering of respondents, can use our method to create their own weights. The creation of such weights is feasible when there is a clearly defined sampling population from which the data set is derived, and when a large number of relevant variables that influence participation probabilities are available in both the study sample and a data set that is representative of the sampling population from which the study sample is drawn. However, correcting for volunteer bias using IP weighting is no panacea for highly selected samples (see below).

Some other studies have attempted to control for selection bias into the UKB by constructing IP weights [14, 20]. We also know of one attempt to construct IP weights for a subset of the UKB (namely, the cohort that was also neurologically imaged) [21]. However, we are the first to calibrate such weights on microdata from the UK Census. As such, our study distinguishes itself from prior attempts in at least two ways. First, the use of rich UK Census microdata allows us to include many more variables, and their interactions, into the construction of our weights. The large sample size of the 5% UK Census subsample that we use further adds to the precision of the estimated weights. Second, and importantly, our IP weights make the UKB more representative of the UKB-eligible population, rather than the population of Great Britain. We achieve this by restricting the UK Census data based on age range, region of residence and the location of the 22 assessment centers. Restricting the data set on which the UKB weights are calibrated to the UKB-eligible population only (rather than the full population of Great Britain) is of key methodological importance, as a central assumption to the validity of IP weights estimation is the assumption of sufficient overlap between the sample and the target population (i.e., all conditional probabilities 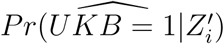 must be different from zero) [19]. This assumption is no longer satisfied when the target population includes types of individuals that are observed in the UKB with probability zero. This would occur, for example if we were to include people living outside the sampling range from any assessment center (e.g., the North of Scotland), or who are outside the sampled age range.

There are some limitations that researchers who wish to use IP weighting in their own association studies need to keep in mind. These limitations also pertain to the results presented here. Our proposed method of IP weighted regression reduces volunteer bias, but may increase standard errors. We suggest therefore that practitioners use our IP weights in UKB analyses as a robustness check, to assess whether their results are not driven by volunteer bias. Further, a main limitation of our IP weighted analyses is the possibility of misspecification of the equation that is used to estimate the likelihood of UKB participation. Although we have used a functional form that is as flexible as possible, given the data, and have separated our data into a training and testing sample to prevent overfitting, we are limited to the inclusion of a limited number of variables that the UKB and UK Census have in common. These are, by and large, variables that measure differences between individuals along a sociodemographic and health dimension. Our method therefore mostly corrects for socioeconomic status, health and demographic-related components of study participation. There may exist unobserved variables that nonetheless explain a substantial part of UKB participation, e.g., personality characteristics. Although our model includes self-reported health as an input, this variable is unlikely to capture the full range of health characteristics that influence study participation. Such unobserved variables could lead to misspecification of the UKB participation model. As a result, any associations estimated in the UKB that correct for volunteer bias may still not be reflective of the true associations in the underlying UKB-eligible population.

This possibility for misspecification is the main reason why collecting representative data is always preferable over an attempt to correct associations estimated in non-representative data using IP weighting. Nonetheless, keeping these drawbacks in mind, our results presented in Subsection 2.4, and in the accompanying Supplementary Note S3, suggest our weights reduce a substantial part of selection bias in UKB-estimated associations. Hence, our IP weights are a reasonable alternative to assessing selection bias in associations estimated in the UKB.

## 4 Methods

### 4.1 Data

#### 4.1.1 UK Biobank (UKB)

The UKB is a cohort of 502,500 individuals collected between 2006 and 2010 at 22 assessment centers spread out across Great Britain [22]. Potential participants were identified through the registry of the National Health Service, which covers virtually the whole population of Great Britain. Individuals living in proximity to an assessment center and aged 40 to 69 at the start of the assessment period (which varies per assessment center) received an invitation to participate by post. These assessment centers were disproportionately located in highly urbanized areas (see Figure 2 and Table S1). This UKB-eligible population consists of 9,238,453 individuals who received an invite, such that the overall acceptance rate was 5.45%.

We drop individuals who died before UK Census day (March 27th) and individuals with missing relevant variables. Next, we remove individuals who do not meet the criteria for the UKB-eligible population. We drop all participants who were not aged between 40 and 69 at the start of their assessment center’s sampling period. That is, for assessment centers that started sampling in 2007, we keep all individuals born between 1937 and 1967, for assessment centers that started sampling in 2008, we keep all individuals born between 1938 and 1968, etc. However, we keep individuals born in 1969 assessed at the Bristol center (which started assessment in 2008), and keep individuals born in 1970 assessed at the Birmingham center (which started assessment in 2009), as these centers both sampled a significant number of individuals born in these years.

All variables that we use were assessed through a touchscreen questionnaire at assessment, or were derived from the NHS registry. We harmonize the coding of some variables to ensure that their categorization is similar to UK Census-derived variables (see Supplementary Note S1).

#### 4.1.2 UKB: Geographic sampling

To ensure that the subsample derived from the UK Census (described in Section 4.1.3) is representative of the UKB-eligible population, we need to understand to which geographic areas, around each of the 22 assessment centers, UKB invitations were sent. Figure S4 shows the place of residency of all UKB participants on the day they visited the assessment center. Other authors have claimed that UKB assessment centers sent out invites to all individuals in the targeted age range that were living within a radius of 40 km [4]. However, a closer assessment of the data reveals that, for virtually all assessment centers, this sampling radius is considerably smaller, and the size of the sampling area varies per assessment center. For example, Figure S4 shows that the assessment center in Edinburgh only sampled respondents in relative proximity (max 22.4 km), whereas the assessment center in Middlesbrough sent out invitations to a wider area (max 39.9 km).

We obtain the sampling radius of each assessment center from the UKB data as follows: for each assessment center, we assume that the UKB participant who lived furthest away was also the furthest living person to receive an invite for that center, and we define the assessment center’s sampling radius accordingly.^5^ However, this method is sensitive to outliers. For 14 assessment centers, we obtain sampling radii that are unrealistically large, as the difference between the furthest participant and the participant in the 99.9th percentile of the distance-to-assessment center distribution is more than 2 km. For these centers, we define the sampling radius as the 99.7th percentile of distances to the assessment center, plus 2 km. The resulting sampling radii are visualized as the circles in Figure S4. A small number of UKB participants (108) fall outside the sampling radii of any assessment center, and are excluded from our data set.

To ensure that our UKB participation probabilities are robustly estimated, we dropped UKB participants residing in Census grouped local authority (GLA) districts with fewer than 70 UKB participants. This resulted in a small loss of another 263 UKB respondents. The final UKB dataset used in our main analyses includes 491,268 UKB participants.

#### 4.1.3 UK Census data

The 2011 Census Microdata Individual Safeguarded Samples (Local Authority) for England and Wales [23], and Scotland [24] are a random 5% subsample of the 2011 UK Census (*N* ≈ 3.1 million). The UK Census collects a wide range of demographic variables at the individual and household level every 10 years through a paper-based or online questionnaire, and covers the whole UK. 2011 was the Census year closest to the UKB assessment period. We restrict the Census data to observations that would have been eligible for sampling into the UKB during its data collection period from 2006 to 2010 (the “UKB-eligible population”), using information on their year of birth and region of residence. First, we only keep individuals aged 40 to 74 at the time of the Census, as these could have been aged between 40 and 69 between 2006 and 2010. However, the cutoff values of the UKB age distribution vary by assessment center, as different assessment centers sampled during different years between 2006 and 2010. Hence, we restrict the age range of Census observations further as discussed below.We use the age of the individual at the day of the Census (5-year bins) to infer the year of birth bin of each individual.

Second, we infer exactly which regions of residence UKB included in its sampling population, and only restrict the UK Census data to include respondents living in these regions. Location of residence in the UK Census data is reported at a higher level of aggregation than in the UKB, namely by *grouped local authority* (GLA) *region* (grouped council authority; GCA, for Scotland, named GLA here for convenience). These regions consist of a single local authority when the population in these regions was larger than 120,000, and of aggregated groups of neighboring local authorities otherwise. There are 285 distinct GLA regions covering all of Great Britain. We keep only those Census individuals that resided in a GLA region that falls *at least partially* within the sampling radius of one of the 22 assessment centers in England, Wales, or Scotland (see Figure S4).

Restricting the UK Census based on age and region as described above introduces some individuals in our sample that may not have received a UKB invite during the period 2006 to 2010, because they were not in the relevant age range (40 to 69) at the time that their nearest assessment center started sampling, or because they lived too far from a UKB assessment center. We calculate adjustment factors to ensure that our UK Census sample is as representative as possible of the UKB-eligible population in 2006 and 2010, conditional on survival up until March 2011. First, we assign each individual in our UK Census data set an initial adjustment factor of 20, to upweight the 5% random subsample available to us to the full UK population. We next adjust these adjustment factors in the following two ways.

First, individuals who resided in GLA regions that did not fully fall within the UKB sampling radius of any assessment center have their adjustment factor multiplied by the proportion of the population living within this GLA region that are within the sampling radius. These GLA-specific sampling population proportions are calculated using 2011 UK Census population counts reported for the much less aggregated *lower layer super output areas* (LSOAs) for England and Wales. For Scotland, even less aggregated *output areas* (OAs) are used because LSOAs are unavailable. For England and Wales, there are 34,753 distinct LSOAs and for Scotland, there are 46,351 OAs. Figure S4 shows the GLA regions that are included in the final sample, and illustrates how adjustment factors are assigned to each Census respondent living in these regions (grey scales range from adjustment factors of 0 [white] to 20 [black] and anything in between [grey]).

Second, the year of birth distribution of the UKB is assessment-center specific, due to the fact that some assessment centers started sampling sooner than others. For example, the assessment center in Manchester sampled all its respondents in 2007, and hence only sampled respondents born between 1937 and 1967, as these were aged 40 to 69 at the time. By contrast, the center in Swansea sampled all its respondents in 2010, and hence only sampled respondents born between 1940 and 1970. We aim to create a one-to-one mapping between the year of birth distributions in the UKB and the UK Census, but face the extra challenge that the UK Census only reports year of birth bins of 5 years. As a result, the year of birth distribution of our UKB-eligible UK Census sample and the UKB do not necessarily overlap, i.e., not everyone in the year of birth bins 1936-1940 or 1966-1970 (at the edges of the cohort distribution) was UKB-eligible. For Census individuals in these year of birth bins, we assign them to an assessment centre based on the GLA in which they live. We multiply their adjustment factor by the proportion of year of birth values (out of a maximum of five) in this year of birth bin that were sampled by the assessment centre. For example, Census respondents residing in or around Manchester that were born between 1936-1940 have their sampling weight multiplied by 0.8, since Manchester only sampled those with year of birth 1937, 1938, 1939, and 1940, but not 1936. When a GLA region overlaps with the sampling radius of multiple assessment centers, we adjust conservatively by taking the largest possible adjustment factor out of the multiple assessment centres with which this GLA overlaps.

The final UKB-eligible census subsample that we use consists of 687,491 observations. Our adjustment factors for this sample range between 0.22 and 20 with a mean of 15.8 and a median of 18.6. This implies a UKB-eligible population of 10,836,059 individuals (as given by the sum of all these sampling weights), slightly larger than the true number of invitations that were send out by the UKB (9,238,452).

Throughout, we regard statistics derived from this UKB-eligible UK Census subsample as unbiased with respect to the UKB’s sampling population. Thus, we assume that the UK Census microdata is itself representative of the UK population and does not suffer from any selection biases. This assumption seems realistic since the UK Census aims to survey the whole UK population, had a response rate of over 95% [23], and the UK Census variables we use have few missing observations (see Supplementary Note S2). Furthermore, the goal of the UK Census is to obtain representative statistics for the UK population. We harmonize the coding of some variables to ensure optimal comparability between these and UKB-derived variables (see Supplementary Note S1).

### 4.2 Inverse probability weighting

Inverse probability weighting (IPW) is a method to correct for volunteer bias in observational data [19, 25, 26]. We model the likelihood of participating in the UKB for individual *i*, conditional on having received an invitation, 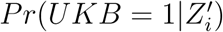, as

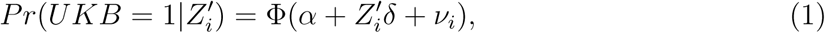

with Φ(.) the standard normal cumulative distribution function (CDF), *α* a constant, *ν*_*i*_ a random error term, and 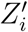 a vector of variables that influences one’s individual propensity for participating in the UKB. Variables included in 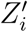 are sex, year of birth (5-year cohort), education level, ethnicity, region of residence (Census GLA), tenure of dwelling, employment status, number of cars in the household, a dummy indicating whether the person lives in a single-person household, and self-reported health. These variables are included in a non-parametric manner (i.e., we use dummy variables for each category of the categorical variables under consideration). Furthermore, we include all possible two-way interactions between these dummy variables as predictors. In total, 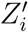 contains 4,820 variables.

For estimation, we only consider the UKB-eligible UK Census sample, constructed as described in Subsection 4.1.3. Estimation is conducted on a randomly selected training sample of 80% UKB-eligible Census data and 20% UKB data. All UKB participants included in the training sample therefore represent 4 sampled individuals from the UKB-eligible population, and hence are assigned an adjustment factor of 4 to give proportional weight to the UKB-eligible UK Census and the UKB data in the estimation of the IP weights used to correct for volunteering bias. In estimation, the UKB-eligible UK Census sample is weighted according to their sampling weights as described in Subsection 4.1.3

We estimate Equation 1 by weighted probit regression on the training sample of stacked UKB and UKB-eligible UK Census data, where we assign the outcome variable *UKB* = 1 to each UKB observation, and *UKB* = 0 to each UKB-eligible UK Census observation. To prevent overfitting, we estimate the model using a LASSO variable selection procedure [27]. The LASSO model maximizes the log-likelihood function of the regular probit, subject to the absolute value of the sum of the coefficients being smaller than a certain constant (as determined by a penalization parameter *λ*). This additional constraint in the optimization problem prevents overfitting of the data when many regressors are included, as it ensures that coefficients of variables that are insufficiently predictive of selection are being shrunk to zero. We estimate our LASSO probit model using *glmnet* [28], which solves the optimization problem

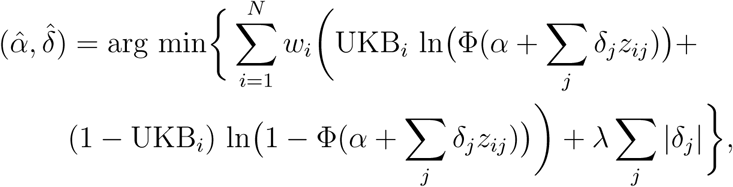

where *w*_*i*_ is the weight (4 for UKB observations, and the adjustment factor constructed as described in Subsection 4.1.3 for UK Census observations), and *λ* is the penalization parameter.

The penalization parameter is chosen through cross-validation using k-folding with 5 folds. The k-folding procedure ensures that *λ* is chosen as to yield an optimally predictive model on a holdout sample not used in the estimation of the LASSO model. This results in a penalization parameter of 0.00017. 2,113 out of the 4,680 variables we include had their coefficients shrunk to zero. This low penalization parameter implies that the solutions to our model lie very close to those of a similarly specified regular probit model in which the same variables (including the twoway interactions) are included.

From the resulting LASSO probit predictions, we next construct inverse probability weights as

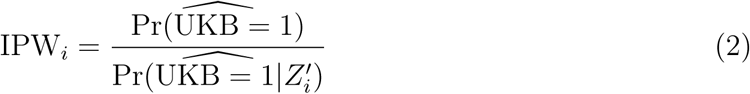

where 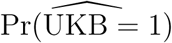 is the average probability of being sampled in the UKB as estimated on the full weighted stacked UKB and UKB-eligible UK Census data set, and 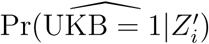 is the probability of UKB participation for UKB participant *i* as predicted by the LASSO probit model.

A known issue with the estimation of inverse probability weights using a rich set of predictors is that such rich models may yield some values of 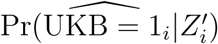 that are very close to zero, with excessively large values of IPW_*i*_ as a result. Such excessive weights can result in noisy estimates of weighted regression coefficients and hence dilute power [19]. We deal with this issue by winsorizing our distribution of estimated weights, setting any values of IPW_*i*_ lower than the 1st percentile equal to the value at the first percentile, and any values of IPW_*i*_ higher than the 99th percentile equal to the value at the 99th percentile.

### 4.3 Weighted association analyses

We use our IP weights to correct various statistics in the UKB for volunteer bias. Weighted means 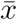 for each variable *x* are constructed by 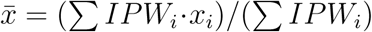. Weighted standard deviations are constructed by 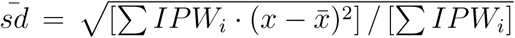. Correspondingly, the 95% confidence interval for the weighted mean is given by 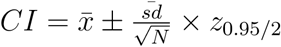.

In Subsection 2.4, we use our IP weights to correct various regressions estimated in the UKB for volunteer bias. These models are estimated by weighted least squares. Intuitively, this weighted regression puts a higher weight on UKB participants that were less likely to participate in the UKB, and a lower weight on participants that were more likely to participate. As a result, estimated regression coefficients from these weighted regressions are more representative of what these regression coefficients would have looked like, if we could have performed the regression on the UKB-eligible population, rather than on the subset of the UKB-eligible population that decided to participate. The method assumes that a participant of the UKB that is assigned IPW_*i*_ is truly representative of non-sampled individuals with the same probability to participate in the UKB, in terms of the exposure and outcome. Furthermore, all conditional probabilities 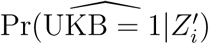 must be different from zero, an assumption which is satisfied as long as the distributions of the conditional probabilities in the UKB and the UKB-eligible population overlap.

P-values that test the null hypothesis that estimates in the UKB and the UKB-eligible UK Census data are the same are calculated using the following Z-statistic: 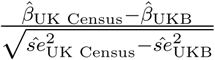

Last, we characterize the *absolute* bias for each model, where a model is typically a linear association between two variables, as follows. First, we estimate the bias of the UKB-based estimate (not IP weighted) as the absolute difference between the UKB-eligible UK Census point estimate 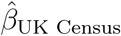 and the UKB point estimate 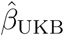. These differences provide us with a measure of volunteering bias for each model in the UKB. Second, we apply IP weights *IPW*_*i*_ to the UKB. After IP weighting, the bias is recalculated by taking, once more, the absolute difference between the (now IP weighted) UKB point estimates 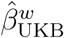 and 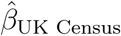. To obtain an overall assessment of volunteering bias across all models (i.e. all associations), we take the mean absolute bias of unweighted UKB estimates, minus the mean absolute bias of the weighted UKB estimates, and divide by the mean absolute bias of unweighted UKB estimates. We find that IP weighting reduces volunteering bias by about 78%.

## Data Availability

The present study uses UK Census safeguarded microdata and UK Biobank data. UK Census safeguarded microdata data and UKB data can be accessed upon request for research projects that have obtained necessary approval. These requests can be submitted through https://ukdataservice.ac.uk/ and https://www.ukbiobank.ac.uk/ respectively.

https://ukdataservice.ac.uk/

## Acknowledgements

Research reported in this publication was supported by the National Institute On Aging of the National Institutes of Health (RF1055654 and R56AG058726), the Dutch National Science Foundation (016.VIDI.185.044), and the Jacobs Foundation. This research has been conducted using the UK Biobank Resource under Application Number 55154. We thank participants at the 2021 BGA annual meeting, 2021 ASHG conference, and the 2021 Integrating Genetics and Social Science Conference for their feedback and comments.

## Supplementary Material

### S1 Harmonization of variable categories across UKB and UK Census

To estimate our IPWs, estimate our summary statistics in Subsection 2.2 and to estimate the regressions in Subsection 2.3 and Subsection 2.4, we selected variables that were assessed in a similar fashion in the UKB and the UK Census. When needed, we altered the categorization of several variables in the UKB and UK Census to ensure that the variables were comparable across both data sets.

#### S1.1 Year of Birth

- **UK Census:** Year of birth was derived from 5-year age bins that described the age of the individual at the day of the UK Census (40-44; 45-49; 50-54; 55-59; 60-64; 65-69; and 70-74). These were recoded into the following year of birth values: (1966-1970; 1961-1965; 1956-1960; 1951-1955; 1946-1950; 1941-1945; 1936-1940).^6^
- **UKB:** Year of birth in the UKB was assessed from the NHS registry. UKB respondents had the possibility to correct their year of birth if it was wrong. We recoded year of birth into the 5 year of birth bins mentioned above.

#### S1.2 Sex

- **UK Census:** Respondents were asked “What is your sex?”, and could answer male or female.
- **UKB:** Sex as recorded by the NHS registry, but possibly updated by the participant.

#### S1.3 Region of residence

- **UK Census:** Region of residence was inferred from the respondent’s address. The respondent’s address was prefilled by the Census data collectors, and corrected by the respondent when necessary. Region of residence is reported as 265 distinct grouped local authorities (GLA; England & Wales), and 20 grouped council areas (GCA; Scotland). GLAs or GCAs consist of local authorities and council areas, or groups of adjacent local authorities and council areas to ensure that each GLA/GCA has at least 120,000 inhabitants.
- **UKB:** Coordinates of home location at assessment, inferred from NHS registry data on the post code level, rounded to the nearest kilometer. These coordinates were aggregated into the same GLAs and GCAs mentioned above, using .shp files that describe the borders of these GLAs and GCAs, obtained from the office for national statistics.

#### S1.4 Ethnicity

- **UK Census:** Respondents were asked “What is your ethnic group?”, and could respond “White”, “Mixed”, “Asian/Asian Birtish”, “Black/African/Caribbean/Black British”, or “Other”
- **UKB:** Wording of the question was the same as in the UK Census. The available answer categories were also the same, except that “Chinese” was recognized as a seperate ethnic group. We merged this group with “Asian/Asian British”. Additionally, respondents could answer “None of the above” or “Prefer not to answer”. We coded these responses as missing.

#### S1.5 Economic status

- **UK Census:** UK Census respondents were asked what their main economic activity was last week through various questions. Answers were coded by the Census bureau into the following categories:

1. Economically Active (excluding Full-time students), In Employment, Employee, Part-time
2. Economically Active (excluding Full-time students), In Employment, Employee, Full-time
3. Economically Active (excluding Full-time students), In Employment, Self employed with employees, Part-time
4. Economically Active (excluding Full-time students), In Employment, Self employed with employees, Full-time
5. Economically Active (excluding Full-time students), In Employment, Self employed without employees, Part-time
6. Economically Active (excluding Full-time students), In Employment, Self employed without employees, Full-time
7. Economically Active (excluding Full-time students), Seeking work and ready to start within 2 weeks, and Waiting to start a job already obtained and available to start within 2 weeks
8. Economically Active Full-time students, In employment
9. Economically Active Full-time students, unemployed, seeking work and ready to start within 2 weeks, and waiting to start a job already obtained and available to start within 2 weeks
10. Economically Inactive, Retired
11. Economically Inactive, Student
12. Economically Inactive, Looking after home/family
13. Economically Inactive, Permanently sick/disabled
14. Economically Inactive, Other

We recoded these levels into a sparser number of categories, namely “Employed” (1-6, 8), “Retired” (10), “Stay-at-home” (12), “Incapacitated” (13), “Unemployed” (7), and “Student” (9, 11)

- **UKB:** At baseline, UKB respondents were asked in the touchscreen questionnaire: “Which of the following describes your current situation?”. Participants could respond to this question as follows:

1. In paid employment or self-employed
2. Retired
3. Looking after home and/or family
4. Unable to work because of sickness or disability
5. Unemployed
6. Doing unpaid or voluntary work
7. Full or part-time student

-7 None of the above

-3 Prefer not to answer

We classified those doing unpaid or voluntary work as “Stay-at-home”, and coded those answering “None of the above” or “Prefer not to answer” as missing.

#### S1.6 Tenure of household

- **UK Census:** Respondents answered the question: “Does your household own or rent this accommodation?” They could answer that they own it outright, own it with a mortgage or loan, part own and part rent (shared ownership), rent, or live rent-free.
- **UKB:** Respondents answered the question: “Do you own or rent the accommodation that you live in?” If repondents tapped the “Help” button they were shown the following: “Please select: - Own outright if you or someone in your household owns the accommodation that you live in. - Own with mortagage if you or someone in your household has a mortagage on the accommodation that you live in.” Answers were the same as in the UK Census, except that respondents could also answer “None of the above”, or “Prefer not to answer”. We coded these as missing.

#### S1.7 Number of vehicles in the household

- **UK Census:** Respondents answered the question: “In total, how many cars or vans are owned, or available for use, by members of this household?” (Please include company vehicles if available for private use). They could answer none, 1, 2, 3, or 4 or more.
- **UKB:** The wording of this question was identical to the wording in the UK Census, as were the answering categories. In addition, we coded those answering “None of the above” or “Prefer not to answer” as missing.

#### S1.8 Household size

- **UK Census:** The only variable regarding household size available to us in the UK Census data was a dummy that indicated whether the person resided in a one-person household or not.
- **UKB:** Respondents answered the question;”Including yourself, how many people are living together in your household? (Include those who usually live in the house such as students living away from home during term, partners in the armed forces or professions such as pilots”. We recoded the answers into a dummy variable that described whether the answer was 1, or some higher number. In addition, we coded those answering “None of the above” or “Prefer not to answer” as missing.

#### S1.9 Self-reported health

In the UK Census, respondents answered to the question “How is your health in general?”. In the UKB, respondents answered to the question “In general how would you rate your overall health?”. Self-reported health in the UKB was assessed through a 4-level Likert scale (Poor, Fair, Good, Excellent), whereas self-reported health in the 2011 UK Census was assessed through a 5-level scale (Very Bad, Bad, Fair, Good, Very Good). To minimize misclassification error, we harmonize these values in both data sets to a three-level scale (Bad, Fair, Good/Excellent), by combining the categories “Very Bad” and “Bad” in the UK Census, and lumping Good/Excellent/Very good into a single category. In addition, respondents in the UKB could answer “Do not know” or “Prefer not to answer”, these answers were coded as missing.

#### S1.10 Education

In the UK Census, respondents were asked “Which qualifications do you have?” and were instructed to tick every box that applied. These answers were recoded by the Census bureau into the highest level of education obtained for each respondent. In the UKB, respondents were asked the question “Which of the following qualifications do you have? (You can select more than one)”. However, the potential answers to each question differed between the UK Census and the UKB.

We harmonized the level of education across both data sets as follows: The UK Census data assigned ISCED levels to the variable “highest degree obtained”. We assigned years of education based on these levels: 7 for no degree (assuming primary school), 10 for a level 1 or level 2 degree, 13 for a level 3 degree, and 20 for a level 4+ degree. This assignment follows previous work in the UKB [29]. There were also two other categories in the UK Census that were not assigned any ISCED level in the UK Census data. These categories were “apprenticeship” and “Other: Vocational/Work-related qualifications, etc.”. We assigned 12 years to the “apprenticeship” (reflecting the fact that it requires continuing one’s education after GCSEs, but does not require an A/AS-level degree) [30]. For the “vocational” category, we again followed previous work [29], and assigned 15 years of education.

For the UKB, we similarly assigned years of education to degree categories that clearly fall within the categories recognized by the UK Census. These categories are: “No degree” (ISCED1), “College or university” (ISCED4+), “A levels/AS levels or equivalent” (ISCED3), “O levels/GCSEs or equivalent” (ISCED2) and “CSEs or equivalent” (ISCED2). However, for those who reported having a “NVQ or HND or HNC or equivalent” or “Other professional qualifications, e.g. nursing, teaching”, assigning years of education was not as straightforward. For those holding an NVQ, we do not know which level of NVQ certificate they hold (In the UK Census, an NVQ of level 1 is considered an ISCED1 degree, whereas an NVQ of level 4 or higher is considered ISCED4+). Accordingly, substantial heterogeneity in the variable “age at which left full time education” can be seen for the group of respondents holding a degree in this category, with a substantial group of respondents having left full time education before the age of 16 (Figure S8a). To solve this issue, for those holding an NVQ (or HND or HNC) we assign years of education by taking the age at which the respondent reported to have left full time education, minus 5, and cap the value at 19 years of education. For those holding a professional degree, we similarly observed substantial heterogeneity in the age at which these respondents left full time education (Figure S8b). Hence, for this group, we estimate years of education in similar fashion, but cap the variable at 15 years. When UKB respondents reported multiple degrees, we take the maximum of the years of education associated with each degree.

These continuous years of education measures make educational attainment comparable across the UKB and the UK Census. For estimating the selection model, we discretize the variable. Those less than 8.5 years of education get level 1 (no degree), those between 8.5 and 11 get level 2 (O-levels or equivalent), those between 11 and 17.5 level 3 (A-levels, vocational, or equivalent), and those above 17.5 get level 4 (college/university, or equivalent).

### S2 Missing data in the UK Census and UK Biobank, and imputation procedure

Both the UK Census and UKB had missing data on the variables that we use to predict UKB participation. Table S4 provides an overview for each variable that we use. In the UKB, respondents explicitly had the option to not share information on all variables measured through self-reporting. They could either tick the options “prefer not to answer” or “do not know”. We coded such values as missing. As a result, 4.9% of our included UKB respondents had missing data on at least one predictor included in the selection model. In the UK Census, data on the variables that we use is typically not missing, but for some variables regarding the household in which the individuals live (i.e., tenure of dwelling, number of cars owned, and household size), no information is available for 0.63% of the observations, as these were people living in communal establishments.

Our model uses a large number of regressors to predict UKB selection status. In the training and holdout samples combined, 24,380 UKB and 4,355 UK Census observations had at least one regressor missing. These missing values were imputed using an exact matching procedure. We conduct exact matching by converting the data to a frame that holds the following variables: whether it was a UK Census or UKB data point, region, year of birth, sex, education, self-reported health, employment status, and sex. In step one, we fill in missing values by sampling from observations with the exact same values on all these variables. For observations for which an exact match could not be found, we attempt to match again using the same variables, but dropping region and ethnicity from the dataframe. For 60 observations, this procedure did not yield an exact match, such that IP weights could not be estimated.

### S3 Robustness of IP weighted regressions to missing variables and miscoding

We haven taken great care in harmonizing variables across the UKB and the UK Census. Nonetheless, subtle differences in the phrasing of questions or the phrasing of answers between these two data sets could introduce artificial differences in the distribution of these variables between the UKB and UK Census that are due to reasons other than selection bias. As a result, our LASSO probit model may not exclusively pick up changes between the UKB and UK Census that are due to selection bias, which introduces bias in our IP weights. To understand whether our IPWs are robust against such misspecification bias (and also missing variable bias), we perform a robustness check in which we re-estimate our weights based on a sparser set of variables.

We choose three sets of variables on which to re-estimate our LASSO probit model, and consecutively our IPWs, and next compare each new set of IP weighted regressions to the results based on weights that use the full set of variables. The three sets of variables that we choose are as follows.

- Set 1 includes only variables that are objectively well-measured in the UKB and UK Census, namely, sex, year of birth, and region of residence.
- Set 2 includes these variables, and additionally also includes ethnicity, and socioeconomic variables that were measured in a highly similar way across both data sets: tenure of dwelling, number of cars, employment status, and the one-person household dummy.
- Set 3 includes all these variables, and self-reported health. Self-reported health was measured in a similar fashion across both data sets, but using a 4-level likert scale in the UK Census and a 5-level likert scale in the UKB. This change in the detail of self-reported health assessment may introduce some unexpected differences between both data sets.
- Finally, we include the original set with all variables, which includes all of the variables in set 3, and our educational attainment measure. Educational attainment was assessed quite differently in both data sets, which might have resulted in some inconsistency across both data sets.

Figure S6 shows the same regressions as reported in Figure 3, but adding the weighted regressions using IPWs based on sparser variables. The sparse model with only three variables captures very little of the selection bias in the UKB, and bias only gets reduced by 9%. This suggests that IPWs based on such a sparse set of variables suffer from omitted variables, and hence these weights are largely unsuccessful at controlling for volunteer bias. Including various socioeconomic variables improves the ability of the IP weights to reduce bias, with a total bias reduction of 59%. This suggests that the addition of self-reported health or education (the variables about which we have the most doubts that they measure the same concepts across the UK Census and the UKB) is not a necessity for the weights to work reasonably. Nonetheless, inclusion of self-reported health does lead to an increase in bias reduction (69%) overall. Inclusion of education improves things a bit more, leading to a total bias reduction of 78% across all models.

Based on these sensitivity analyses, we conclude that all of the variables we used are important to improving the performance of the LASSO probit weights, and therefore that the IP weights based on all variables performs best. No addition of any specific variable that may have been miscoded, or that is not sufficiently comparable across the two datasets, seems to radically alter the estimated associations from the IP weighted regressions in the UKB. This provides confidence in our method.

In Figure S7, we perform the same test for the regressions with discrete/continuous variables, and similar conclusions hold, leading to bias reductions of 4%, 26%, 63%, and 77% for each set of variables, respectively.

**Table S1:** Comparison of the means of various variables in the full population of Great Britain (40-74 year olds) and the UKB-eligible population (40-74) year olds as estimated in the UK Census data. 95% confidence intervals around each mean are included. In the right column, means and standard deviations (used for the confidence intervals) were weighted using adjustment factors that make the UK Census representative of the UKB-eligible population.

| Variable | Mean in full population [95% CI] | Mean in UKB-eligible population [95% CI] |
| --- | --- | --- |
| *Demographic* |  |  |
| Age | **55.098** [55.082;55.115] | **54.784** [54.761; 54.808] |
| Living in a couple | **0.719** [0.718;0.72] | **0.689** [0.688; 0.69] |
| Born outside UK | **0.119** [0.119;0.12] | **0.164** [0.163; 0.165] |
| White | **0.916** [0.916;0.917] | **0.87** [0.869; 0.871] |
| Sex | **1.509** [1.508;1.51] | **1.508** [1.507; 1.509] |
| Household size | **2.61** [2.608;2.613] | **2.65** [2.647; 2.653] |
| *Socioeconomic status* |  |  |
| Deprived in education dimension | **0.264** [0.264;0.265] | **0.264** [0.263; 0.265] |
| Deprived in employment dimension | **0.154** [0.153;0.155] | **0.179** [0.178; 0.18] |
| Deprived in health and disability dimension | **0.357** [0.356;0.358] | **0.371** [0.369; 0.372] |
| Deprived in housing dimension | **0.082** [0.082;0.083] | **0.098** [0.098; 0.099] |
| Deprivation indicator (total) | **1.857** [1.856;1.859] | **1.912** [1.909; 1.914] |
| Years of education | **12.652** [12.643;12.66] | **12.638** [12.625; 12.652] |
| Owns house | **0.407** [0.406;0.408] | **0.362** [0.36; 0.363] |
| No. of cars | **1.463** [1.462;1.465] | **1.366** [1.363; 1.368] |
| *Health* |  |  |
| Self-reported health | **3.96** [3.958;3.962] | **3.91** [3.908; 3.913] |
| Disability | **0.235** [0.234;0.236] | **0.247** [0.246; 0.248] |
| Number of housecarers in household | **0.335** [0.334;0.336] | **0.346** [0.344; 0.348] |
| No. in household with illness/disability | **0.449** [0.448;0.45] | **0.468** [0.466; 0.47] |
| *Employment* |  |  |
| Employed | **0.612** [0.611;0.613] | **0.613** [0.612; 0.614] |
| Retired | **0.222** [0.222;0.223] | **0.217** [0.215; 0.218] |
| Unemployed | **0.053** [0.053;0.053] | **0.04** [0.039; 0.04] |
| Ever worked | **0.81** [0.809;0.811] | **0.866** [0.864; 0.867] |
| *Urbanicity* |  |  |
| Persons per room | **0.407** [0.407;0.407] | **0.424** [0.424; 0.425] |
| Goes to work by public transport | **0.113** [0.112;0.114] | **0.167** [0.165; 0.168] |
| Goes to work by car/motorcycle | **0.642** [0.641;0.643] | **0.635** [0.633; 0.637] |
| Lives in flat or apartment | **0.135** [0.134;0.135] | **0.163** [0.162; 0.164] |
| *Religion* |  |  |
| Has no religion | **0.2** [0.2;0.201] | **0.182** [0.181; 0.183] |
| Christian | **0.674** [0.673;0.674] | **0.667** [0.666; 0.668] |
| Other religion | **0.126** [0.125;0.127] | **0.151** [0.15; 0.152] |
| Observations | 1277785 | 566502 |

**Table S2:** Full summary statistics for the UKB-eligible population and the UKB. The UKB-eligible population is created by restricting and weighting (using adjustment factors) the 5% subsample of the 2011 UK Census. 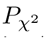 in the 7th column reports the p-value of a *χ*^2^-test for equal distributions for each variable as measured in the UKB-eligible UK Census population and the UKB.

| Variable | Levels | Census (UKB-eligible) | | UKB | | $P_{\chi^2}$ |
| --- | --- | --- | --- | --- | --- | --- |
|  |  | N | % | N | % |  |
| Sex | Male | 5,333,695.06 | 49.22 | 222,971 | 45.39 | < 1e-8 |
|  | Female | 5,502,364.27 | 50.78 | 268,297 | 54.61 |  |
|  | Total | 10,836,059.32 | 100.00 | 491,268 | 100.00 |  |
| Birthyear | 1936-1940 | 674,012.26 | 6.22 | 33,586 | 6.84 | < 1e-8 |
|  | 1941-1945 | 1,289,775.68 | 11.90 | 101,081 | 20.58 |  |
|  | 1946-1950 | 1,671,907.91 | 15.43 | 114,996 | 23.41 |  |
|  | 1951-1955 | 1,624,169.90 | 14.99 | 82,433 | 16.78 |  |
|  | 1956-1960 | 1,874,898.22 | 17.30 | 70,732 | 14.40 |  |
|  | 1961-1965 | 2,151,457.40 | 19.85 | 61,117 | 12.44 |  |
|  | 1966-1970 | 1,549,837.96 | 14.30 | 27,323 | 5.56 |  |
|  | Total | 10,836,059.32 | 100.00 | 491,268 | 100.00 |  |
| Education | None | 2,920,933.24 | 26.96 | 83,553 | 17.01 | < 1e-8 |
|  | Lower secondary | 2,832,405.03 | 26.14 | 141,037 | 28.71 |  |
|  | A-levels/vocational | 2,069,303.74 | 19.10 | 94,150 | 19.16 |  |
|  | University | 3,013,416.20 | 27.81 | 161,511 | 32.88 |  |
|  | Total | 10,836,058.20 | 100.00 | 480,251 | 100.00 |  |
| Ethnicity | White | 9,625,885.91 | 88.83 | 464,757 | 94.60 | < 1e-8 |
|  | Mixed | 105,552.61 | 0.97 | 2,891 | 0.59 |  |
|  | Asian/Asian British | 656,703.51 | 6.06 | 11,273 | 2.29 |  |
|  | Black/Black British | 356,016.68 | 3.29 | 7,879 | 1.60 |  |
|  | Other | 91,900.62 | 0.85 | 4,468 | 0.91 |  |
|  | Total | 10,836,059.32 | 100.00 | 491,268 | 100.00 |  |
| Health (self-reported) | Bad | 1,003,746.72 | 9.26 | 21,711 | 4.42 | < 1e-8 |
|  | Fair | 2,054,220.11 | 18.96 | 102,875 | 20.94 |  |
|  | Good/Very Good | 7,778,091.38 | 71.78 | 364,370 | 74.17 |  |
|  | Total | 10,836,058.20 | 100.00 | 488,956 | 100.00 |  |
| Employment status | Paid employment | 6,602,507.40 | 60.93 | 281,812 | 57.36 | < 1e-8 |
|  | Retired | 2,693,524.53 | 24.86 | 165,944 | 33.78 |  |
|  | Stay-at-home | 385,636.64 | 3.56 | 13,607 | 2.77 |  |
|  | Incapacitated | 751,258.09 | 6.93 | 16,066 | 3.27 |  |
|  | Unemployed | 359,497.74 | 3.32 | 7,998 | 1.63 |  |
|  | Student | 43,634.92 | 0.40 | 1,284 | 0.26 |  |
|  | Total | 10,836,059.32 | 100.00 | 486,711 | 100.00 |  |
| No. of vehicles in household | 0 | 1,992,849.34 | 18.50 | 42,717 | 8.70 | < 1e-8 |
|  | 1 | 4,350,708.42 | 40.40 | 204,076 | 41.54 |  |
|  | 2 | 3,243,366.45 | 30.12 | 185,911 | 37.84 |  |
|  | 3 | 881,116.19 | 8.18 | 41,968 | 8.54 |  |
|  | 4 or more | 301,482.86 | 2.80 | 13,160 | 2.68 |  |
|  | Total | 10,769,523.25 | 100.00 | 487,832 | 100.00 |  |
| Tenure of dwelling | Owns house (no mortgage) | 3,812,493.95 | 35.40 | 254,890 | 51.88 | < 1e-8 |
|  | Owns house w/ mortgage | 4,109,126.28 | 38.16 | 180,172 | 36.67 |  |
|  | Shared ownership | 49,412.74 | 0.46 | 1,428 | 0.29 |  |
|  | Rent | 2,707,334.15 | 25.14 | 44,185 | 8.99 |  |
|  | Rent-free | 91,156.12 | 0.85 | 3,482 | 0.71 |  |
|  | Total | 10,769,523.25 | 100.00 | 484,157 | 100.00 |  |
| One-person household | No | 8,822,644.06 | 81.92 | 397,792 | 80.97 | < 1e-8 |
|  | Yes | 1,946,879.19 | 18.08 | 90,130 | 18.35 |  |
|  | Total | 10,769,523.25 | 100.00 | 487,922 | 100.00 |  |

**Table S3:** Means of various variables in the UKB before and after weighting using IPWs: 95% confidence intervals around each mean are included.

| Variable | Mean [95% CI] | Weighted Mean [95% CI] | % Change |
| --- | --- | --- | --- |
| *Anthropometric* |  |  |  |
| Height | **168.713** [168.684;168.741] | **169.148** [169.119; 169.178] | 0.3% |
| BMI | **27.415** [27.4;27.429] | **27.645** [27.63; 27.661] | 0.8% |
| Waist Circumference | **90.319** [90.277;90.361] | **90.881** [90.838; 90.924] | 0.6% |
| Waist Hip Ratio | **0.871** [0.871;0.872] | **0.876** [0.875; 0.876] | 0.5% |
| Hip Circumference | **103.451** [103.423;103.479] | **103.574** [103.544; 103.603] | 0.1% |
| *Demographic* |  |  |  |
| Age at Recruitment | **56.54** [56.518;56.563] | **53.603** [53.579; 53.627] | 5.2% |
| Urban | **0.862** [0.861;0.863] | **0.875** [0.874; 0.875] | 1.5% |
| Died after Census Day | **0.035** [0.034;0.035] | **0.036** [0.035; 0.036] | 2% |
| *Early lifetime* |  |  |  |
| Breastfed | **0.723** [0.722;0.725] | **0.684** [0.683; 0.686] | 5.4% |
| Multiple Birth | **0.023** [0.022;0.023] | **0.024** [0.024; 0.025] | 6% |
| Birth Weight | **3.319** [3.316;3.321] | **3.312** [3.31; 3.315] | 0.2% |
| Adopted as a child | **0.015** [0.014;0.015] | **0.016** [0.016; 0.017] | 11.7% |
| Maternal smoking | **0.292** [0.291;0.294] | **0.297** [0.295; 0.298] | 1.5% |
| *Food/Beverage consumption* |  |  |  |
| Tea | **3.411** [3.403;3.419] | **3.406** [3.397; 3.414] | 0.1% |
| Bread Consumed | **0.849** [0.846;0.852] | **0.836** [0.834; 0.839] | 1.5% |
| Cooked Veg. Consumption | **2.723** [2.718;2.729] | **2.69** [2.684; 2.696] | 1.2% |
| Cheese Consumption | **2.523** [2.52;2.526] | **2.454** [2.45; 2.457] | 2.7% |
| *Health* |  |  |  |
| Disability | **0.289** [0.288;0.29] | **0.301** [0.299; 0.302] | 4% |
| Asthma | **0.127** [0.125;0.129] | **0.134** [0.132; 0.136] | 5.6% |
| DBP | **82.259** [82.225;82.293] | **82.037** [82.002; 82.071] | 0.3% |
| SBP | **140.21** [140.148;140.273] | **138.362** [138.3; 138.424] | 1.3% |
| Vitamin D | **48.695** [48.632;48.757] | **46.239** [46.176; 46.302] | 5% |
| Calcium | **2.38** [2.38;2.38] | **2.378** [2.378; 2.379] | 0.1% |
| Cholesterol | **5.696** [5.693;5.699] | **5.598** [5.595; 5.601] | 1.7% |
| White Blood Cell Count | **6.882** [6.876;6.888] | **7.008** [7.002; 7.014] | 1.8% |
| Red Blood Cell Count | **4.517** [4.516;4.518] | **4.545** [4.544; 4.547] | 0.6% |
| Chest Pain | **0.162** [0.161;0.164] | **0.186** [0.185; 0.187] | 14.7% |
| Hand grip strength (left) | **29.535** [29.503;29.567] | **30.084** [30.051; 30.117] | 1.9% |
| Hand grip strength (right) | **31.672** [31.64;31.703] | **32.204** [32.171; 32.237] | 1.7% |
| *Health behavior* |  |  |  |
| Ever Smoked | **0.602** [0.6;0.603] | **0.608** [0.607; 0.61] | 1.1% |
| Alcohol Freq. | **4.143** [4.139;4.148] | **3.984** [3.98; 3.989] | 3.8% |
| Number of Cigarettes | **18.331** [18.274;18.387] | **18.709** [18.651; 18.768] | 2.1% |
| Ever Addicated | **0.06** [0.059;0.061] | **0.074** [0.072; 0.075] | 22.7% |
| Ever Smoked Cannabis | **0.442** [0.437;0.447] | **0.543** [0.537; 0.548] | 22.7% |
| Max Freq. Cannabis use | **1.656** [1.645;1.666] | **1.732** [1.72; 1.743] | 4.6% |
| *Mental Health* |  |  |  |
| Depression | **0.277** [0.274;0.28] | **0.297** [0.294; 0.3] | 7.2% |
| *Other* |  |  |  |
| Left Handed | **0.093** [0.093;0.094] | **0.095** [0.094; 0.096] | 1.6% |
| Ambidextrous | **0.017** [0.017;0.018] | **0.019** [0.019; 0.019] | 10.2% |
| Happiness | **4.575** [4.571;4.579] | **4.52** [4.516; 4.525] | 1.2% |
| *Socioeconomic* |  |  |  |
| Townsend Cont. | **-1.317** [-1.326;-1.308] | **-0.432** [-0.442; -0.423] | 67.2% |
| No. of people in household | **2.437** [2.433;2.441] | **2.599** [2.595; 2.604] | 6.7% |
| Age completed full-time educ. | **16.718** [16.71;16.726] | **16.727** [16.718; 16.735] | 0.1% |
| Time employed current job | **12.933** [12.894;12.973] | **11.928** [11.89; 11.965] | 7.8% |
| Length working week | **35.256** [35.209;35.304] | **36.147** [36.1; 36.194] | 2.5% |
| Heavy manual work | **1.552** [1.548;1.555] | **1.642** [1.638; 1.645] | 5.8% |
| Household income | **44830.202** [44731.179;44929.225] | **43605.497** [43505.706; 43705.288] | 2.7% |

**Table S4:** Prevalence of missing observations in UK Census (UKB-eligible subsample) and UK Biobank

| **Variable** | **UK Census** | **UKB** |
| --- | --- | --- |
| Sex | 0% | 0% |
| Education | 0% | 2.24% |
| Region | 0% | 0% |
| Year of Birth | 0% | 0% |
| Health (Self-reported) | 0% | 0.47% |
| Tenure of dwelling | 0.63% | 1.45% |
| Employment status | 0% | 0.93% |
| Number of cars | 0.63% | 0.7% |
| One-person household | 0.63% | 0.68% |
| Ethnicity | 0% | 0% |

**Figure S1:**
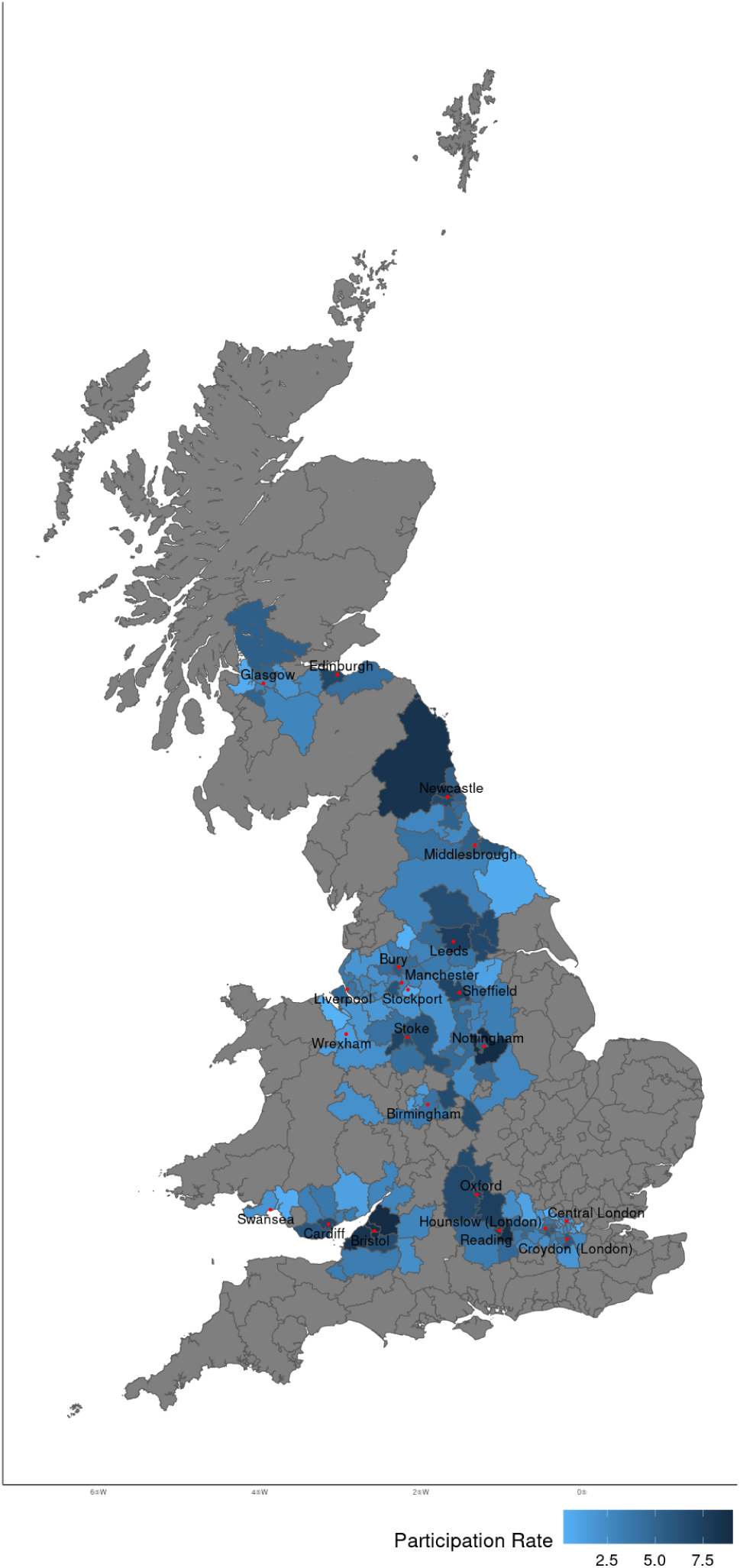
UKB participation rates by region (Census Grouped local authority [GLA] regions). Each participation rate is computed by the number of UKB respondents residing in that GLA (at baseline assessment), relative to the estimated size of the UKB-eligible population residing in that GLA. The size of the UKB-eligible population per GLA is estimated by the number of UKB-eligible individuals in the 5% subsample of the UK Census, and multipled by GLA-specific adjustment factors to reflect the fact that not all Census GLAs fully fell within the sampling radius around each assessment center (see Methods, Subsection 4.1). Citizens residing in grey areas were not sampled by UKB.

**Figure S2:**
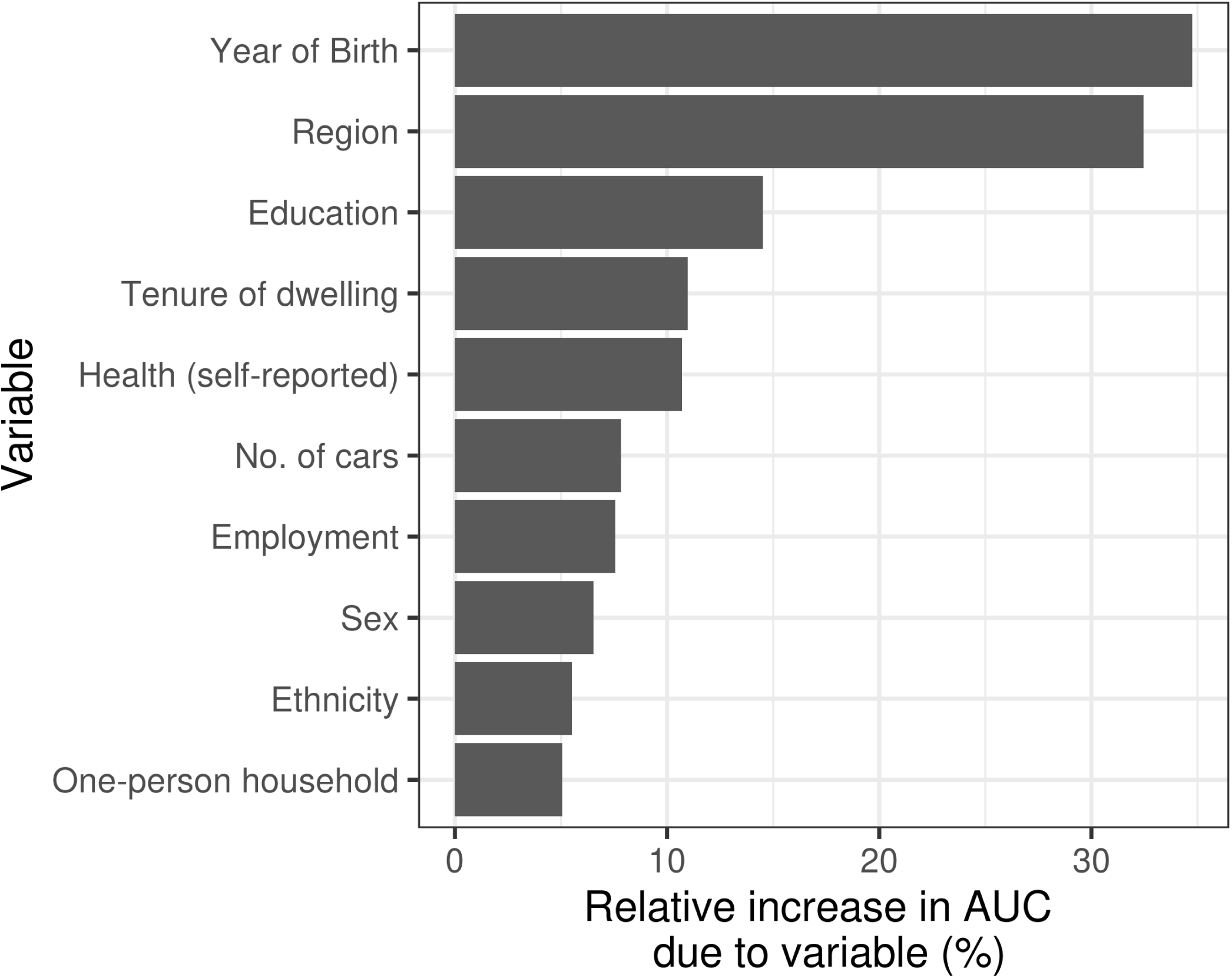
Variable importance plot showing the importance of each variable in predicting UKB volunteering. We quantify the relative increase in AUC due to each variable by permuting each variable in turn. During each permutation, observations for the variable of interest get randomly assigned to individuals in our holdout sample. As a result, the variable can no longer drive any predictive abilities of the model. We next assess the AUC of the model after predicting on the permuted sample. The relative importance of each variable is then quantified as the AUC of the model before permuting the sample, minus the AUC of the model after permuting the sample, divided by the AUC of the model before permuting.

**Figure S3:**
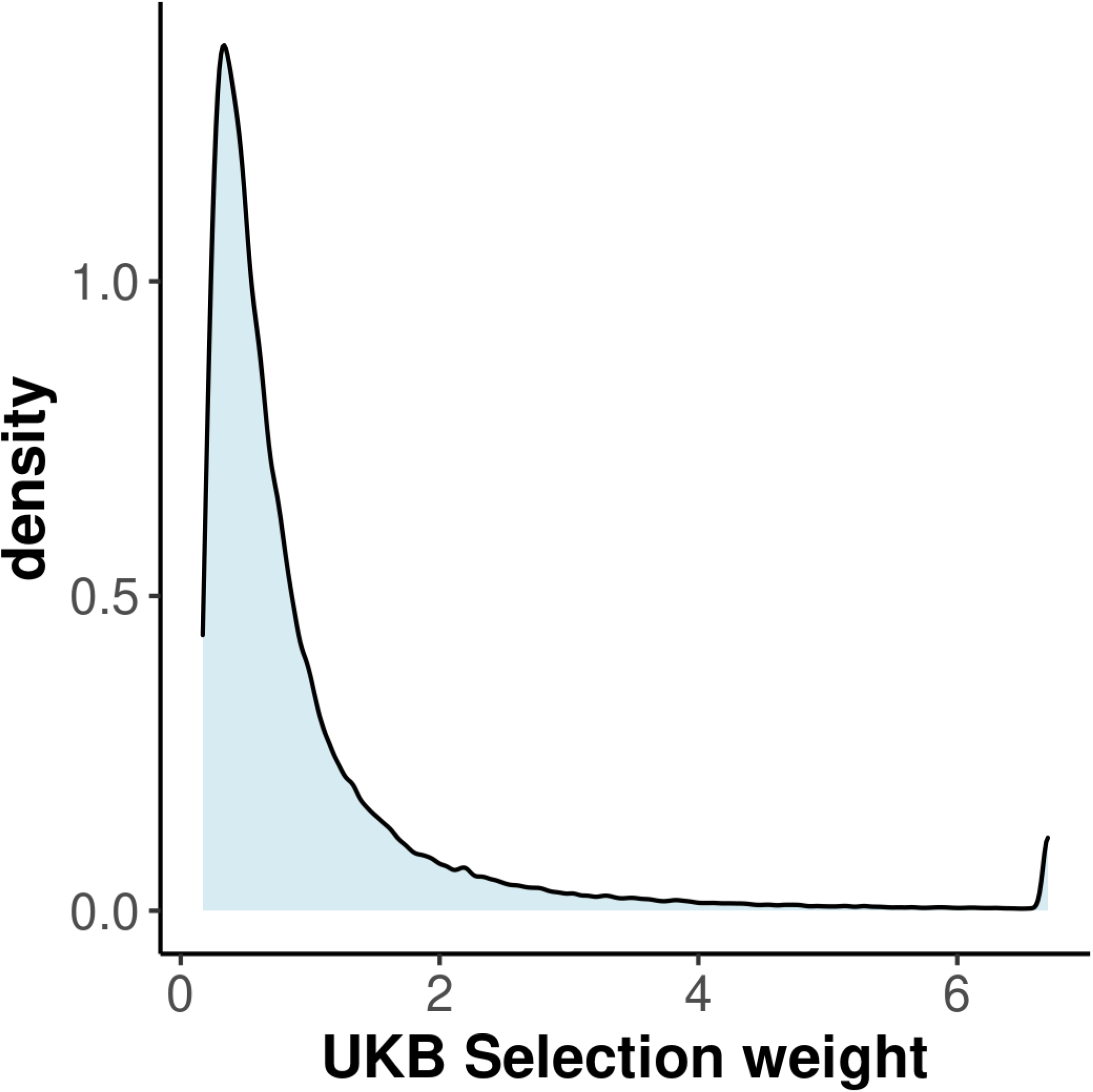
Histogram of the distribution of UKB selection weights. Figure is based on the 80% UKB holdout sample, after winsorizing (setting values below the 1st percentile equal to the value at the 1st percentile, and values above the 99th percentile equal to the 99th percentile.)

**Figure S4:**
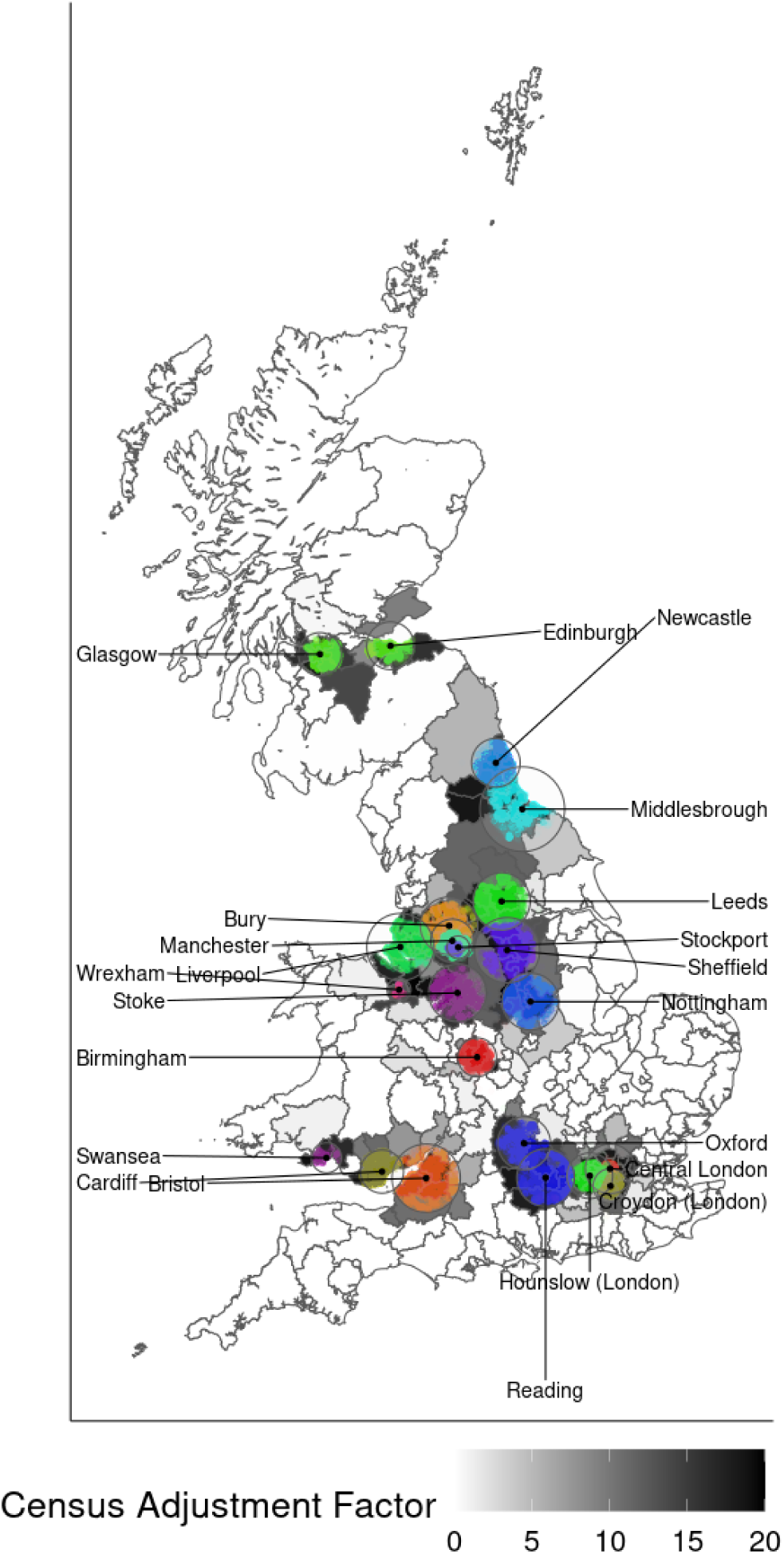
Geographic distribution of UKB participants and the UKB-eligible subsample of the UK Census used to construct UKB participation probabilities. Each colored dot on the map represents the geographic location of a UKB participant’s residence, colored by the assessment center that they visited. Each black dot shows the location of an assessment centre. Each circle visualizes the sampling radius around each assessment center. Census Grouped local authority (GLA) regions that intersect the radius of sampled UKB participants around each assessment center are colored in grey. For GLA regions that are fully within any assessment center’s sampling region, we assign a sampling weight of 20 (darkest grey shades in the map) to each UK Census observation, such that the 5% UK Census subsample that we use becomes representative of the full UKB-eligible population. For Census observations from GLA regions that fall only partially within an assessment center’s sampling region, we use sampling weights (lighter grey shades in the map) of 20 times the share of this region’s population that lives within the assessment center’s radius (see Methods, Section 4.1.3, for details). UKB participants who reside outside any sampling radius (108 in total) were dropped from the data set, as are regions that have no UKB participants (white areas).

**Figure S5:**
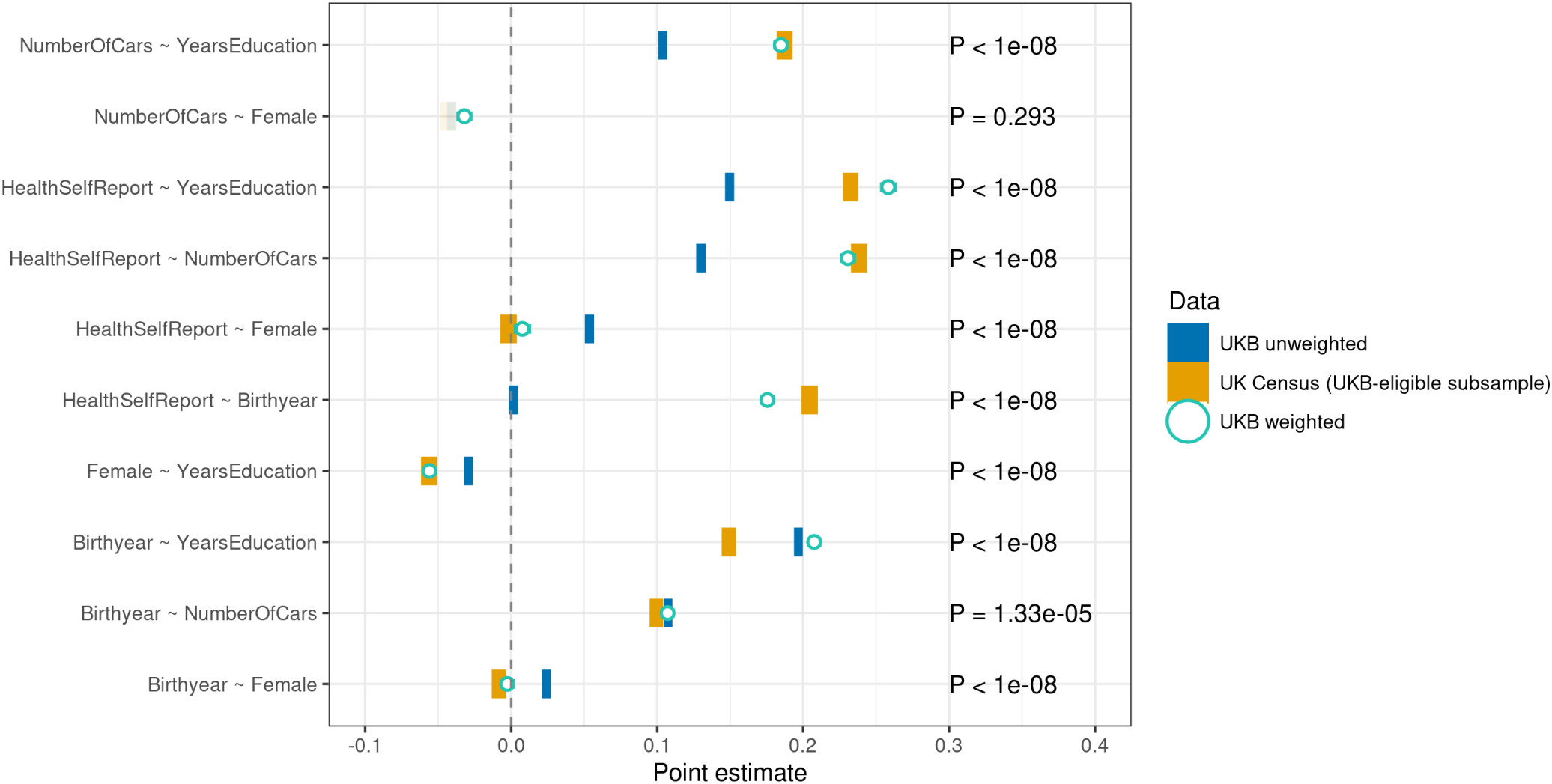
Estimated coefficients for linear models in UKB and UK Census. Each blue bar is estimated in the UKB (80% holdout sample) using OLS. Associations in the UKB-eligible UK Census (yellow bars, 20% holdout sample) are estimated by a WLS model using the UK Census adjustment factors (constructed as described in Methods, Section 4.1.3. The green open circles show WLS models estimated in UKB, with IP weights constructed to correct for volunteering. 95% confidence intervals are indicated by the width of the bar (heteroskedasticity robust standard errors). All variables are standardized to have a mean of zero and a variance of one. All solid blue and yellow bars are significantly different from one another. IP weighting leads to substantially improved associations (closer to the UKB-eligible estimates) with one exception.

**Figure S6:**
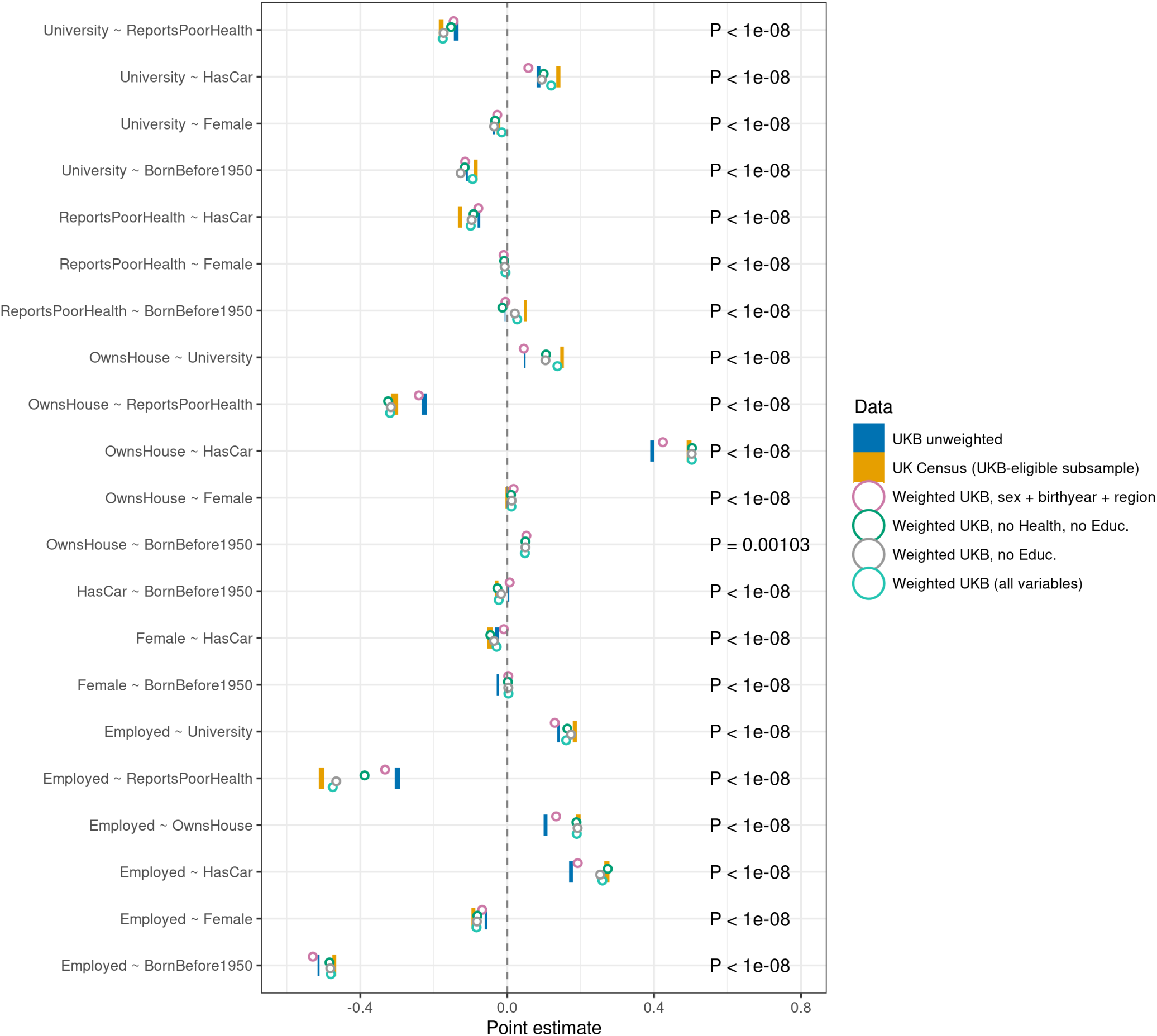
Estimated coefficients for bivariate linear models in UKB and UK Census. Each blue bar is estimated in the UKB (80% holdout sample) using OLS. Associations in the UKB-eligible UK Census (yellow bars, 20% holdout sample) are estimated by a WLS model using the UK Census adjustment factors (constructed as described in Methods, Section 4.1.3. The open circles show WLS models estimated in the UKB, with IP weights constructed to correct for volunteering, based on different sets of variables. 95% confidence intervals are indicated by the width of the bar (heteroskedasticity-robust standard errors). As more variables are included in the LASSO model, IP weighted coefficients move closer and closer to the UKB-eligible subsample (from the blue to the yellow bar).

**Figure S7:**
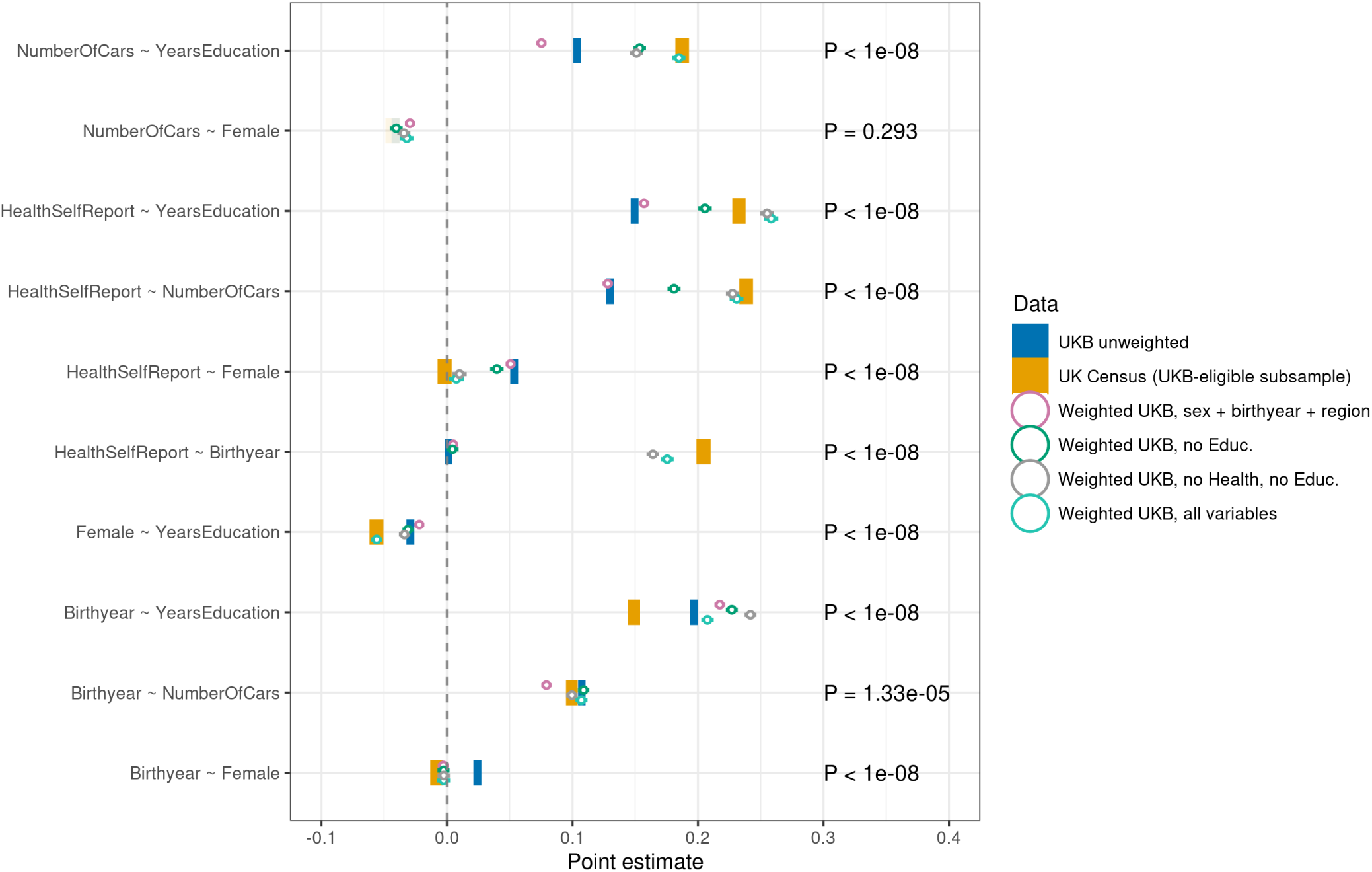
Estimated coefficients for linear models in the UKB and UK Census. Each blue bar is estimated in the UKB (80% holdout sample) using OLS. Associations in the UKB-eligible UK Census (yellow bars, 20% holdout sample) are estimated by a WLS model using the UK Census adjustment factors (constructed as described in Methods, Section 4.1.3). The open circles show WLS models estimated in the UKB, with IP weights constructed to correct for volunteering, based on different sets of variables. 95% confidence intervals are indicated by the width of the bar (heteroskedasticity-robust standard errors). As more variables are included in the LASSO model, IP weighted coefficients generally move closer and closer to the UKB-eligible subsample (from the blue to the yellow bar), but not in all cases.

**Figure S8.**
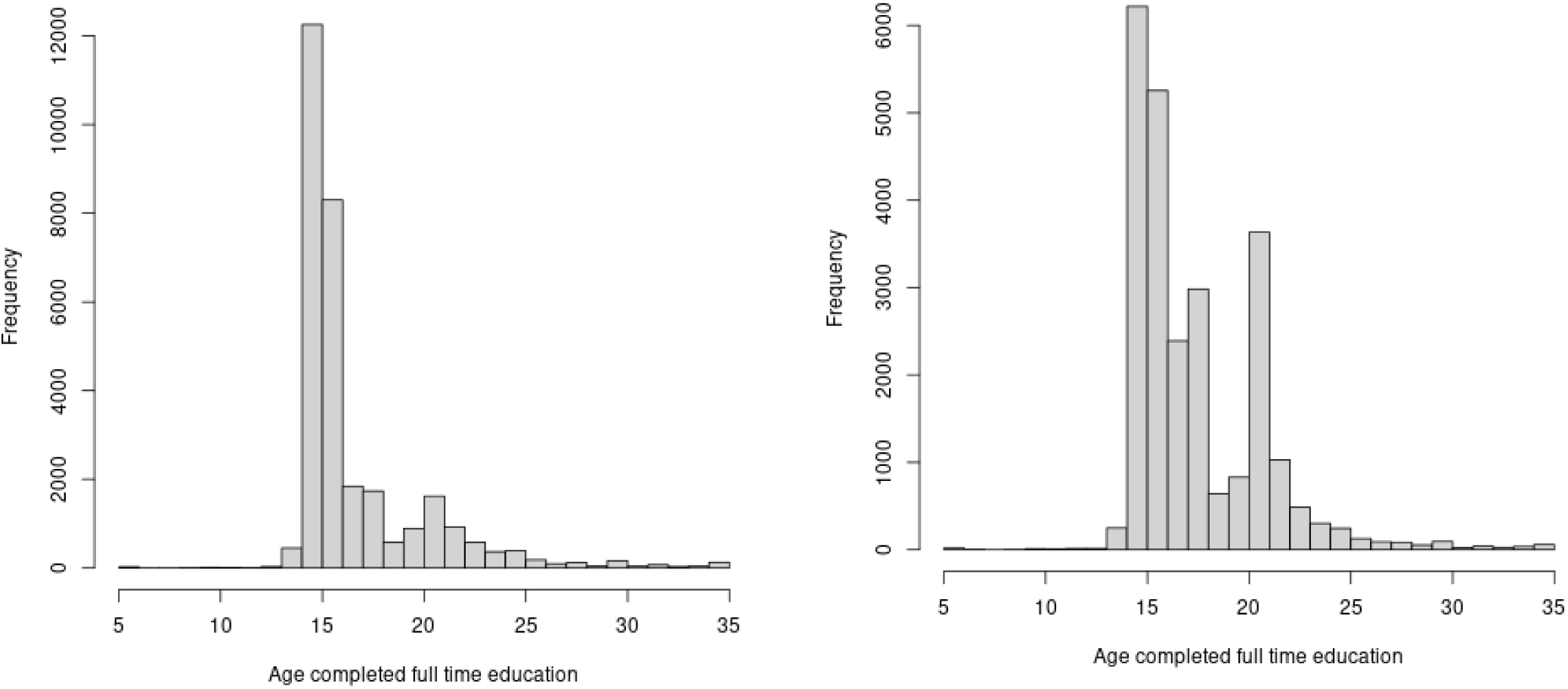
(a) Distribution of “age completed full time education”, for those with an NVQ or HNC or HND or equivalent (b) Distribution of “age completed full time education”, for those with a professional qualification not elsewhere classifed

## Footnotes

1 Northern Ireland was not included in the UKB, nor were any of the smaller British Islands.

2 UKB assessment occurred between 2006 and 2010, with a median assessment date of January 2009. The UK Census was collected before or on UK Census day (27th of March 2011). Hence, there is a time discrepancy between the dates that both data sets were collected. To minimize the consequences of this time discrepancy for our volunteer bias adjustment, we remove from the UKB sample all respondents (N=2,998) who died before UK Census day. Hence, our estimates are also conditional on survival up until the day of the 2011 UK Census.

3 We use 10 categorical variables with 2, 7, 4, 5, 3, 6, 5, 5, 2, and 143 categories respectively. Coding each categorical variable as dummies that reflect the value of each category, and omitting a reference category for each variable, results in 182-10=172 baseline variables. Next, twoway interactions between each of these dummies are included, resulting in (again omitting reference categories) 4,648 interaction terms. Hence, we include a total of 4,648+172=4,820 variables

4 Note that for the bivariate variables, it is not possible to tell whether an increase or a decrease in the standard deviation in the UKB, vis á vis the standard deviation in the UKB-eligible population, is consistent with selective sampling. This is because, for bivariate variables, the standard deviation is 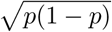, with *p* the mean of the variable. Hence, the standard deviation is largest for *p* = 0.5. When selection into the UKB is such that the mean of the variable becomes closer to 0.5 (from above or from below), the standard deviation will be larger in the UKB than in the UKB-eligible population. For example, this is the case in Table 1 for “University or equivalent”: in the UKB-eligible population, 27.8% holds such a degree, whereas in the UKB, this is 33.6%. This change in the mean of the variable is consistent with selective sampling in the UKB (where healthy and higher educated citizens are more likely to participate in scientific studies), but nonetheless results in a larger standard deviation of this variable in the UKB.

5 We reassign individuals who lived within 40 km of an assessment center, but visited an assessment center further away, to their nearest assessment center.

6 The 2011 UK Census was conducted at 27th of March. We classify each respondent in their 5 year of birth bin assuming that they had *not yet* had their birthday in 2011. This approach inevitably results in some small classification error (e.g., someone who turned 65 on February 1st of 2011 has year of birth 1946, is classified in age bin 65-69 by the UK Census, and is next, erroneously, classified in year-of-birth-bin 1941-1945 by us).

